# Can we use machine learning to discover risk factors? Testing the proof of principle using data on >11,000 predictors and mortality in the UK Biobank

**DOI:** 10.1101/2021.05.07.21256791

**Authors:** Iqbal Madakkatel, Ang Zhou, Mark McDonnell, Elina Hyppönen

## Abstract

**Background:** We present a simple and fast hypothesis-free machine learning pipeline for risk factor discovery that accounts for non-linearity and interaction in large biomedical databases with minimal variable pre-processing.

**Methods:** Mortality models were built using gradient boosting decision trees (GBDT) and important predictors were identified using SHAP values. Cox models controlled for false discovery rate were used for interpretability and further validation. The pipeline was tested using information from 502,506 UK Biobank participants, aged 37–73 years at recruitment and followed over seven years for mortality registrations.

**Results:** From the 11,639 predictors included in GBDT, 193 potential risk factors had SHAP values 0.05 or greater and were selected for further modelling. Of the total variable importance summed up, 60% was directly health related, and baseline characteristics, sociodemographics, and lifestyle factors each contributed about 10%.

Cox models adjusted for baseline characteristics, showed evidence for an association with mortality for 166 out of the 193 predictors. These included mostly well-known risk factors (e,g, age, sex, ethnicity, education, material deprivation, smoking, physical activity, self-rated health, BMI, and many disease outcomes), and the pipeline was able to detect associations in the presence of interactions and non-linear relationships. For 19 predictors we saw evidence for an association in the unadjusted but not adjusted analyses, suggesting bias by confounding..

**Conclusion:** Our GBDT-SHAP pipeline was able to identify relevant predictors ‘hidden’ within thousands of variables, providing a fast and pragmatic solution for the first stage of hypothesis free risk factor identification.

What was known before this study?
Existing studies have used ML for variable selection in general settings, however, we are not aware of studies integrating epidemiological approaches with ML for risk factor discovery in deeply phenotyped large scale population cohorts. We present a simple and fast method for risk factor discovery using GBDT-SHAP pipeline, followed by subsequent standard epidemiological analyses. We test this pipeline using mortality prediction in the UK Biobank as an example.

What this study adds?
Machine learning pipeline integrated with traditional epidemiological practices as described in our study can be used a simple and fast method for discovering potential risk factors in extensively phenotyped high dimensional biomedical databases. GBDT-SHAP pipeline was able to detect associations with factors presenting with interactions with other explanatory variables and non-linear relationships with the outcome with minimal variable pre-processing. However, ML does not replace the requirement for content knowledge and more refined modelling to ensure lack of confounding and accurate interpretation.

## INTRODUCTION

Cohort studies and biobanks available for medical research are growing, both in the number of individuals included and the density of information available for the participants. These large databases hold enormous potential for innovation and provide exciting prospects for hypothesis free risk factor discovery. However, in practice, many research projects use only a set of handpicked predictors for their analyses, due to various limitations. Indeed, traditional epidemiological approaches, such as logistic regression and Cox regression are limited in number of independent variables that can be practically included in a single model. They also require lack of multicollinearity among independent variables, and without careful modelling, by default associations are assumed to be linear with no interactions between the explanatory variables. Further challenges in the multivariate context arise from the treatment of and biases caused by missing information.

Machine learning (ML) algorithms provide some attractive solutions for many of these challenges, and they have been found to be effective in developing predictive models based on large sets of variables. As opposed to a hypothesis-driven approach, ML methods enable predictions based on data only, with no prior assumptions on possible presence of interactions or the shape of the association, simply “learning rules from the data” [1]. In particular, supervised ML methods capture complex interactions and non-linear associations among explanatory variables [2, 3], often resulting in better model performance when subsequently applied to real-world data. There has been great interest in comparing model performance among different ML algorithms [4–7]. ML approaches, gradient boosting decision trees (GBDT), support vector machine, K-nearest neighbors, and artificial neural network have been found to outperform traditional risk scoring systems [4, 5, 8, 9]. Among the strongest approaches is GBDT, which according to a review comparing 13 different state-of-art ML methods, was ranked as the best of all methods in tasks related to predictive analytics [10].

In this study, rather than using ML for building competing predictive models, we test the proof of principle for the ability to discover potential risk factors amongst thousands of predictors by combining GBDT modelling with standard epidemiological practices. We use data from over 11,000 predictors and mortality for over 500,000 participants in the UK Biobank. Our novel analysis pipeline uses GBDT, CatBoost implementation [11] for its inherent capability to handle missing values and a large volume of data, without having to convert variables to any specific format. We screen for potentially interesting predictors using SHAP (SHapley Additive exPlanation) values [12, 13], reflecting variable ‘importance’ to guide the selection of mortality predictors for further epidemiological modelling.

## METHODS

### Participants

The UK Biobank is a cohort of over 500,000 participants recruited between March 13, 2006 and October 1, 2010 through 22 assessment centers across England, Wales and Scotland [14]. Data collection during the baseline assessment covered touch screen questionnaire surveys, face-to-face interviews, and physical measurements, with blood sampling and urine collection for genetic assays and biomarker assessments. Further information on disease outcomes was obtained through record linkages, including mortality statistics from the UK Office of National Statistics, cancer registrations, and hospital episodes statistics.

The outcome variable indicating the mortality status of the participants as of March 1, 2016, was created using the UK Biobank date of death field 40000. In this study, we considered those information that were collected at the baseline assessment, including data obtained using the touchscreen questionnaires and results from clinical examinations. In addition, we included disease codings derived from linkage to cancer registrations and hospital episodes statistics. We removed baseline variables which were recorded for less than 95% of the participants. Information obtained from online follow-up surveys or sub-samples of the cohort were excluded from our analyses due to their low coverage. Supplementary Table 1 lists all the variables included.

The UK Biobank project was approved by the National Information Governance Board for Health and Social Care and North West Multi-center Research Ethics Committee (11/NW/0382). Participants provided electronic consent to use their anonymized data and samples for health-related research, to be recontacted for further sub-studies and for the UK Biobank to access their health-related records [15]. This study was conducted under application number 20175 to the UK Biobank.

### Model development pipeline and statistical analyses

The GBDT-SHAP pipeline is shown in Figure 1. As the data were not sufficiently structured for our analyses, we used a specifically designed software package for UK Biobank, PHESANT (PHEnome Scan Analysis) [16], available in R and ran an automated pre-processing step before developing ML models (Supplementary Methods).

**Figure 1.**
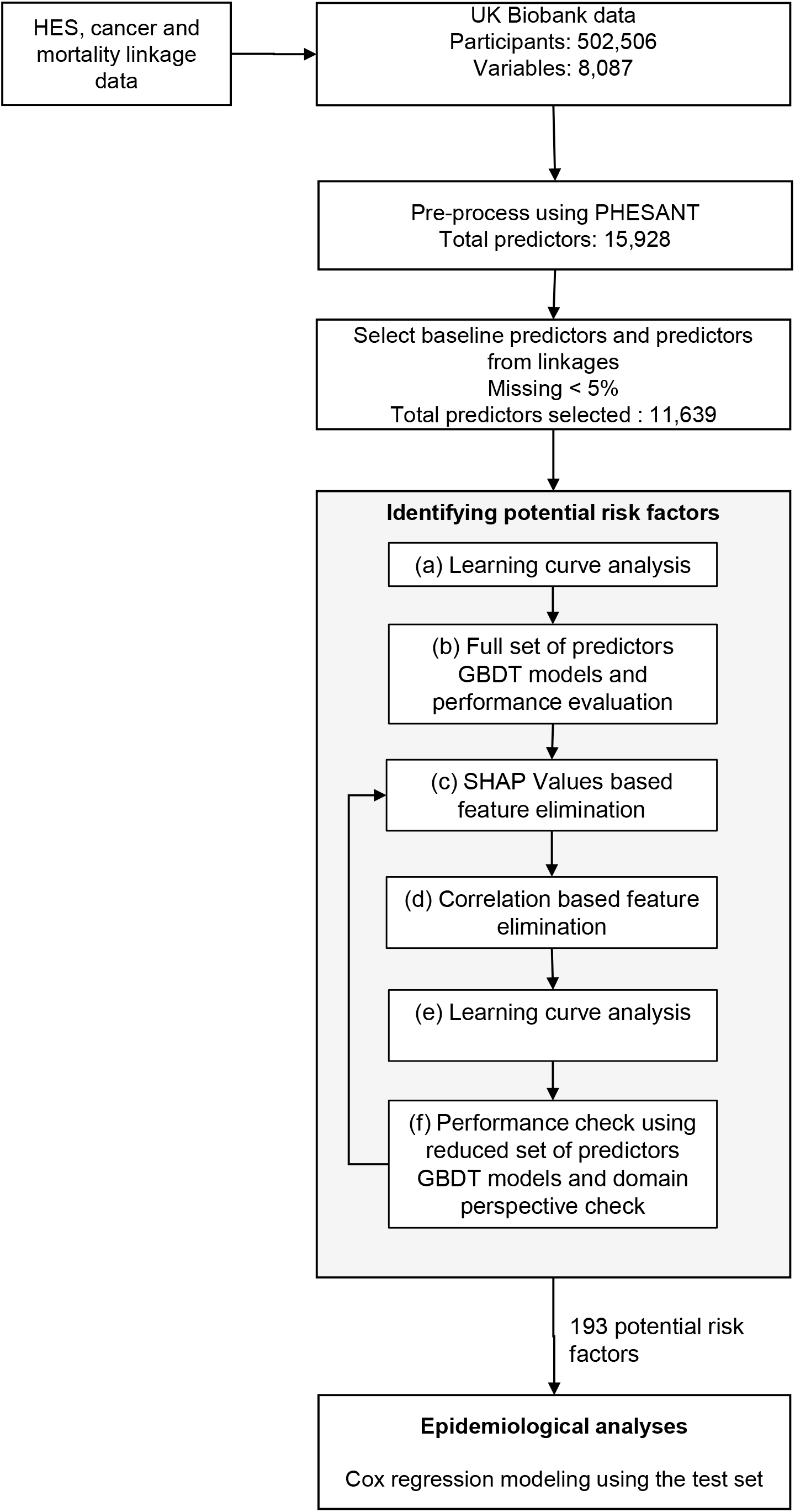
GBDT-SHAP machine learning pipeline for risk factor discovery, followed by epidemiological analyses using Cox regression. GBDT: gradient boosting decision trees; HES: hospital episode statistics; PHESANT: PHEnome Scan Analysis; SHAP: SHapley Additive exPlanation.

### Identifying potential risk factors

Potential risk factors were identified by following the six steps, namely, a) learning curve analysis to determine sufficient amount of data for training b) developing GBDT models with all available predictors and assessing model performance, c) calculating variable importance using SHAP values and eliminating predictors based on a threshold, d) further elimination of highly monotonically correlated predictors, e) learning curve analysis to determine sufficient amount of data for training with the reduced set of predictors and f) ensuring that the reduced set of predictors is appropriate from model performance perspective as well as from an epidemiological perspective.

Our GBDT models used in the above steps are binary classifiers, that is, their input are the predictors for each individual and their output is model’s confidence for mortality status of that individual. The classes were highly imbalanced (death rate was around 2.9%) and to address the class imbalance problem, all our ML models were developed with the hyperparameter ‘positive class weight’ set to the ratio of negative to positive training samples [17, 18]. We initially split the entire data into random training, development, and test sets at the ratio of 60:20:20. The training and the development sets were used as the derivation cohort and the test set as the validation cohort. To avoid overfitting to the training data (which is more common in high-dimensional datasets [19]), we used the development set in all our GBDT models for early stopping of training, and thus effectively tuning the hyper parameter ‘number of estimators’. We assessed model performance using area under the receiver operating characteristics (AUROC), a widely used threshold independent metric in assessing binary classifiers. Confidence intervals of AUROC were calculated using 1,000 bootstrap [20] datasets based on the test set. We also report sensitivity and specificity at Youden index optimal cut-off point [21]. We used CatBoost version 0.21 implemented in Python (Python Software Foundation, version 3.5.2) for GBDT model development.

In step (a), we trained GBDT models with increasing numbers of training samples from the training split, starting from 20,000 participants and incremented by 20,000 each time until all training samples were used and verified the adequacy of training samples. Subsequently, we developed GBDT models with all available predictors and assessed their performance in step (b). In step (c), we calculated the importance of each predictor as the mean absolute SHAP value in the training set and normalized variable importance so that they summed up to 100%. We eliminated ‘irrelevant’ predictors using an arbitrarily chosen SHAP threshold. We explored different thresholds to identify ‘important’ predictors and assessed the effects on model performance when using reduced sets of predictors (step (f)). We used Spearman’s ρ (above 0.9) to identify sets of highly correlated predictors and removed all but one (the one recorded for the greatest number of samples) from those sets to produce the final set of predictors for further epidemiological analyses. Such a step avoided carrying forward predictors such as both ‘left leg fat percentage’ and ‘right leg fat percentage’ for further analyses. We repeated the learning curve analysis (step (e)) and model performance check (step (f)) with the reduced set of predictors GBDT models before proceeding to further analyses.

### Epidemiological analyses

As SHAP values do not provide information on directionality, we conducted epidemiological analyses to allow for direct interpretation of the associations between the predictors and mortality risk. We fitted univariate Cox models, and also adjusted for baseline predictors identified as ‘important’ by GBDT, including age, sex, Townsend deprivation index, assessment center and month of birth using the test dataset. We used FDR to account for multiple testing. We present data as Hazard ratios (HR) and their 95% confidence intervals. For selected known mortality predictors which were picked up by GBDT, but not supported by simple Cox modelling, we fitted non-linear models and accounted for selected interactions. We constructed a loop iterating through two-way interactions between predictors which had shown evidence for an association in the GBDT-SHAP pipeline but not confirmed in the Cox models and other predictors identified by GBDT-SHAP pipeline, to further examine mortality associations. All the interaction analyses in the loop were adjusted for baseline predictors as for the other analyses. As our intention was to test for proof of principle in risk factor discovery rather than predictive modelling, for simplicity, we interpret coefficients from Cox models as ‘average associations’, avoiding the reequipment to test for proportionality of hazards assumption. All epidemiological models were done using STATA (version 15, StataCorp, College Station, TX, USA).

## RESULTS

### Participants characteristics

Of the 502,506 participants included in our study, 14,421 participants (2.9%) died over the median of 7 years (IQR 6.3–7.7 years) of follow-up. Table 1 shows the distribution of the participants as a whole and the control, and the case groups separately along with selected baseline characteristics. The training and development sets had 8,552 and 2,941 deaths respectively, while the test set containing 2,928 deaths. Participants who died during the follow-up were older and more commonly male compared to those who stayed alive. Those who died during the follow-up period were less educated, had poorer self-rated health, were current or previous smokers and from more deprived backgrounds.

**Table 1.**
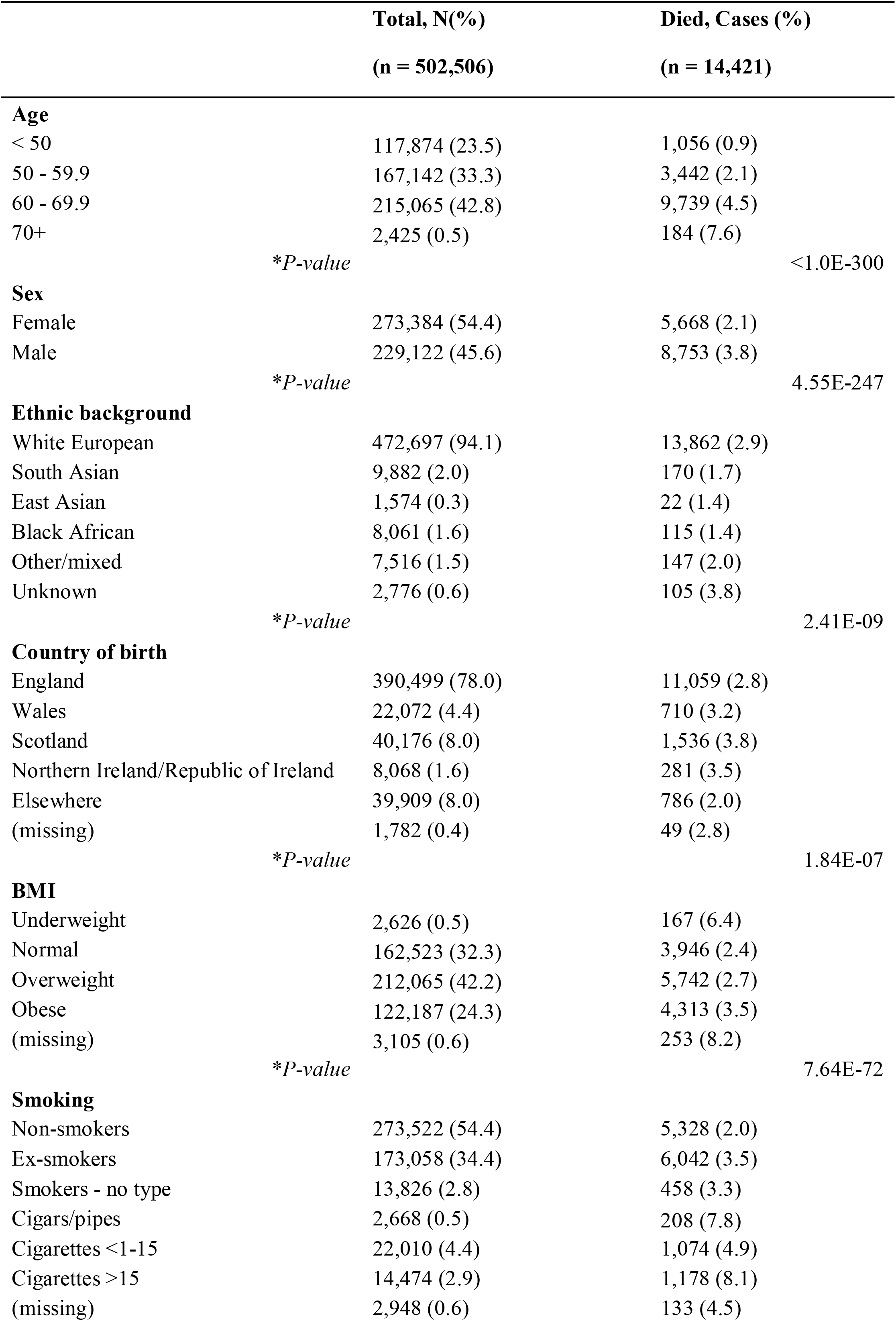
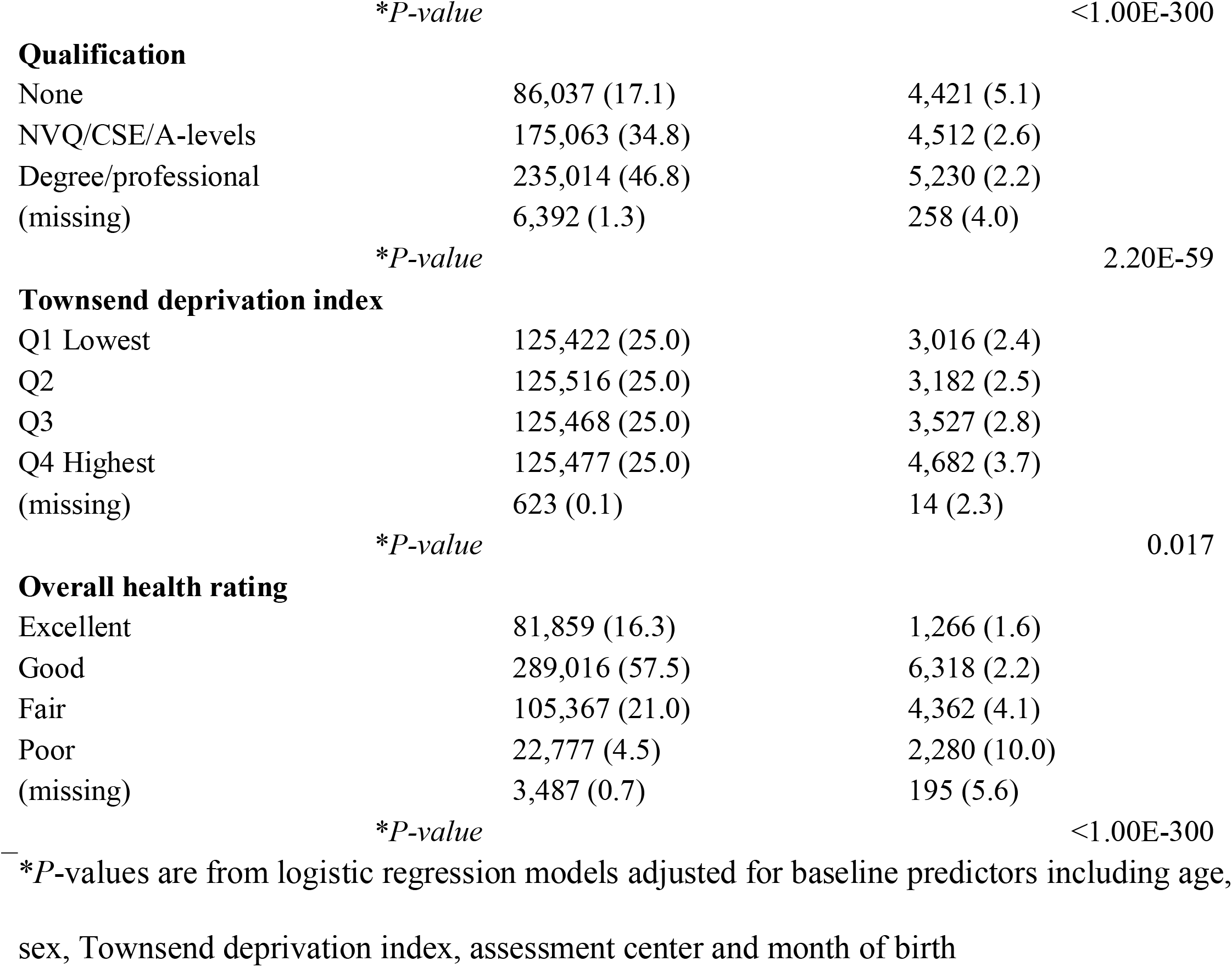
Baseline characteristics of the UK Biobank cohort.

|  | **Total, N(%)**<br>**(n = 502,506)** | **Died, Cases (%)**<br>**(n = 14,421)** |
| --- | --- | --- |
| **Age** |  |  |
| < 50 | 117,874 (23.5) | 1,056 (0.9) |
| 50 - 59.9 | 167,142 (33.3) | 3,442 (2.1) |
| 60 - 69.9 | 215,065 (42.8) | 9,739 (4.5) |
| 70+ | 2,425 (0.5) | 184 (7.6) |
|  | *\*P-value* | <1.0E-300 |
| **Sex** |  |  |
| Female | 273,384 (54.4) | 5,668 (2.1) |
| Male | 229,122 (45.6) | 8,753 (3.8) |
|  | *\*P-value* | 4.55E-247 |
| **Ethnic background** |  |  |
| White European | 472,697 (94.1) | 13,862 (2.9) |
| South Asian | 9,882 (2.0) | 170 (1.7) |
| East Asian | 1,574 (0.3) | 22 (1.4) |
| Black African | 8,061 (1.6) | 115 (1.4) |
| Other/mixed | 7,516 (1.5) | 147 (2.0) |
| Unknown | 2,776 (0.6) | 105 (3.8) |
|  | *\*P-value* | 2.41E-09 |
| **Country of birth** |  |  |
| England | 390,499 (78.0) | 11,059 (2.8) |
| Wales | 22,072 (4.4) | 710 (3.2) |
| Scotland | 40,176 (8.0) | 1,536 (3.8) |
| Northern Ireland/Republic of Ireland | 8,068 (1.6) | 281 (3.5) |
| Elsewhere | 39,909 (8.0) | 786 (2.0) |
| (missing) | 1,782 (0.4) | 49 (2.8) |
|  | *\*P-value* | 1.84E-07 |
| **BMI** |  |  |
| Underweight | 2,626 (0.5) | 167 (6.4) |
| Normal | 162,523 (32.3) | 3,946 (2.4) |
| Overweight | 212,065 (42.2) | 5,742 (2.7) |
| Obese | 122,187 (24.3) | 4,313 (3.5) |
| (missing) | 3,105 (0.6) | 253 (8.2) |
|  | *\*P-value* | 7.64E-72 |
| **Smoking** |  |  |
| Non-smokers | 273,522 (54.4) | 5,328 (2.0) |
| Ex-smokers | 173,058 (34.4) | 6,042 (3.5) |
| Smokers - no type | 13,826 (2.8) | 458 (3.3) |
| Cigars/pipes | 2,668 (0.5) | 208 (7.8) |
| Cigarettes <1-15 | 22,010 (4.4) | 1,074 (4.9) |
| Cigarettes >15 | 14,474 (2.9) | 1,178 (8.1) |
| (missing) | 2,948 (0.6) | 133 (4.5) |
|  | *\*P-value* | <1.00E-300 |
| **Qualification** |  |  |
| None | 86,037 (17.1) | 4,421 (5.1) |
| NVQ/CSE/A-levels | 175,063 (34.8) | 4,512 (2.6) |
| Degree/professional | 235,014 (46.8) | 5,230 (2.2) |
| (missing) | 6,392 (1.3) | 258 (4.0) |
|  | *\*P-value* | 2.20E-59 |
| **Townsend deprivation index** |  |  |
| Q1 Lowest | 125,422 (25.0) | 3,016 (2.4) |
| Q2 | 125,516 (25.0) | 3,182 (2.5) |
| Q3 | 125,468 (25.0) | 3,527 (2.8) |
| Q4 Highest | 125,477 (25.0) | 4,682 (3.7) |
| (missing) | 623 (0.1) | 14 (2.3) |
|  | *\*P-value* | 0.017 |
| **Overall health rating** |  |  |
| Excellent | 81,859 (16.3) | 1,266 (1.6) |
| Good | 289,016 (57.5) | 6,318 (2.2) |
| Fair | 105,367 (21.0) | 4,362 (4.1) |
| Poor | 22,777 (4.5) | 2,280 (10.0) |
| (missing) | 3,487 (0.7) | 195 (5.6) |
|  | *\*P-value* | <1.00E-300 |

— *\*P-values are from logistic regression models adjusted for baseline predictors including age, sex, Townsend deprivation index, assessment center and month of birth*

### Pre-processing

PHESANT pre-processing, after satisfying our missing value criterion, derived 11,639 predictors falling under ten broad categories, baseline characteristics, demographics, lifestyle and environment, physical measurements, cognitive function, psychosocial factors, self-reported diseases, medications and operations, health and medical history and hospital diagnoses (Supplementary Table 2). Hospital diagnoses (through record linkage) accounted for 98% of the predictors.

### Identifying potential risk factors

Our learning curve analysis using all the predictors showed improvements in AUROC as more and more training samples were used (Supplementary Figure 1). We found that in the range of 40% to 60% of samples used for training, model performance stabilized. The GBDT model with all predictors reported an AUROC value of 0.94 (95% CI 0.94 - 0.95) on the test set (Supplementary Figure 2). The model reported a sensitivity of 0.83 and a specificity of 0.92. At an arbitrary cut-off value of 0.05%, 218 predictors were considered to be ‘important’. Correlation based predictor elimination resulted in further reduction of 25 predictors resulting in 193 ‘important’ predictors. Learning curve analysis showed data could be split at the ratio of 60:20:20 also for the reduced set of predictors GBDT models. Reduced set of predictors GBDT model reported an AUROC value of 0.94 (95% CI 0.93 - 0.95). The model had the same sensitivity and specificity as that of all predictors model.

Figure 2 shows the category-wise predictor importance distribution and Supplementary Table 3 lists all important predictors. Hospital diagnoses, health and medical history, and self-reported health jointly covered about 60% of the total variable importance summed up, with baseline characteristics (e.g. age, sex), sociodemographics (e.g. employment, education, housing, ethnicity), and lifestyle factors (e.g. smoking, physical activity, diet) each contributing about 10%. Since the mean absolute SHAP values do not directly indicate the direction of association, we show the SHAP summary plot for all important predictors in Supplementary Figure 3.

**Figure 2.**
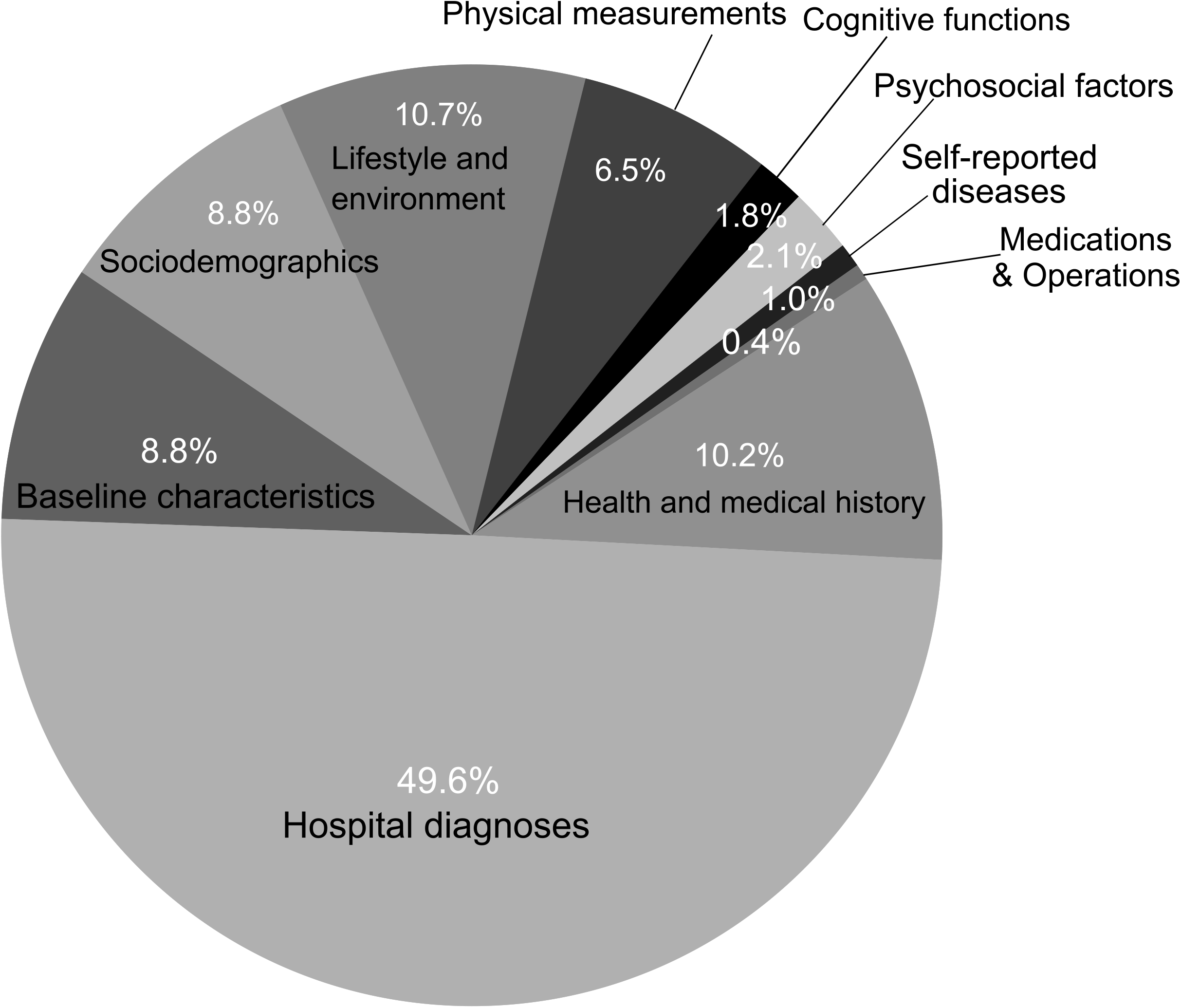
Variable importance values of the 193 important predictors identified for the SHAP value threshold of 0.05%, aggregated into ten categories. Variable importance is calculated as mean absolute SHAP value for each predictor and normalized to 100% before applying the threshold. SHAP: SHapley Additive exPlanation.

### Epidemiological analyses

In Cox models adjusted for age, sex, Townsend deprivation index, assessment center and month of birth, 166 out of 193 predictors had an association with mortality at *P* < 0.05 after correcting for FDR (Supplementary Table 4). Hazard ratios from Cox models of top 50 predictors ranked by SHAP values are shown in Figure 3 and Figure 4. Supplementary Figure 4 shows hazard ratios for all the important predictors. Confirmed predictors included expected mortality associations for various disease outcomes, sociodemographic characteristics and some lifestyle indicators. After FDR correction, there were 19 predictors which showed evidence of association in the unadjusted models but not in the adjusted models, such as length of time at current address, sensitivity/hurt feelings, worrier/anxious feelings, guilty feelings, risk taking, hearing difficulties, whole body fat-free mass, experiencing of headache and knee pain in last month, diagnoses of inguinal hernia, polyp of colon and gonarthrosis. Eight predictors did not meet the *P*-value threshold in either unadjusted or adjusted models, including month of birth, comparative height size at the age of ten, cheese intake, handedness, irritability, using a gas fire in winter time, gastro-esophageal reflux disease without esophagitis, and other and unspecified malignant neoplasm of skin of other and unspecified parts of face. As an attempt to understand why these eight predictors may have been picked up by GBDT-SHAP modelling, we next looked in more detail at their associations with mortality. For example, GBDT picked up month of birth as an important predictor and when we recoded it to seasons, both univariate and multivariate models showed modest evidence for an association (*P* ≤ 0.02). Interaction loop analyses suggested that association between comparative height at age 10 and mortality might have arisen from an interaction with secondary malignant neoplasm of brain and cerebral meninges (*P*_interaction_ = 1.48E-05). Similarly, gastro-esophageal reflux disease without esophagitis showed some evidence for interaction with hypertension (*P*_interaction_ = 0.02) and malignant neoplasm of skin of other and unspecified parts of face had an interaction with fed-up feelings (*P*_interaction_ = 0.009). Other factors such as cheese intake, handedness, irritability, and gas/solid fuel cooking all had low SHAP values (all < 0.08%). Although univariate Cox model showed an association between tea intake and mortality, an adjusted Cox model showed no evidence for a linear association, (*P*= 0.31). However, there was significant non-linearity (*P*_curvature_ = 2.27E-10), with lower mortality for participants drinking 1 to 7 cups per day compared to non-drinkers and the very high intake group (Supplementary Figure 5).

**Figure 3.**
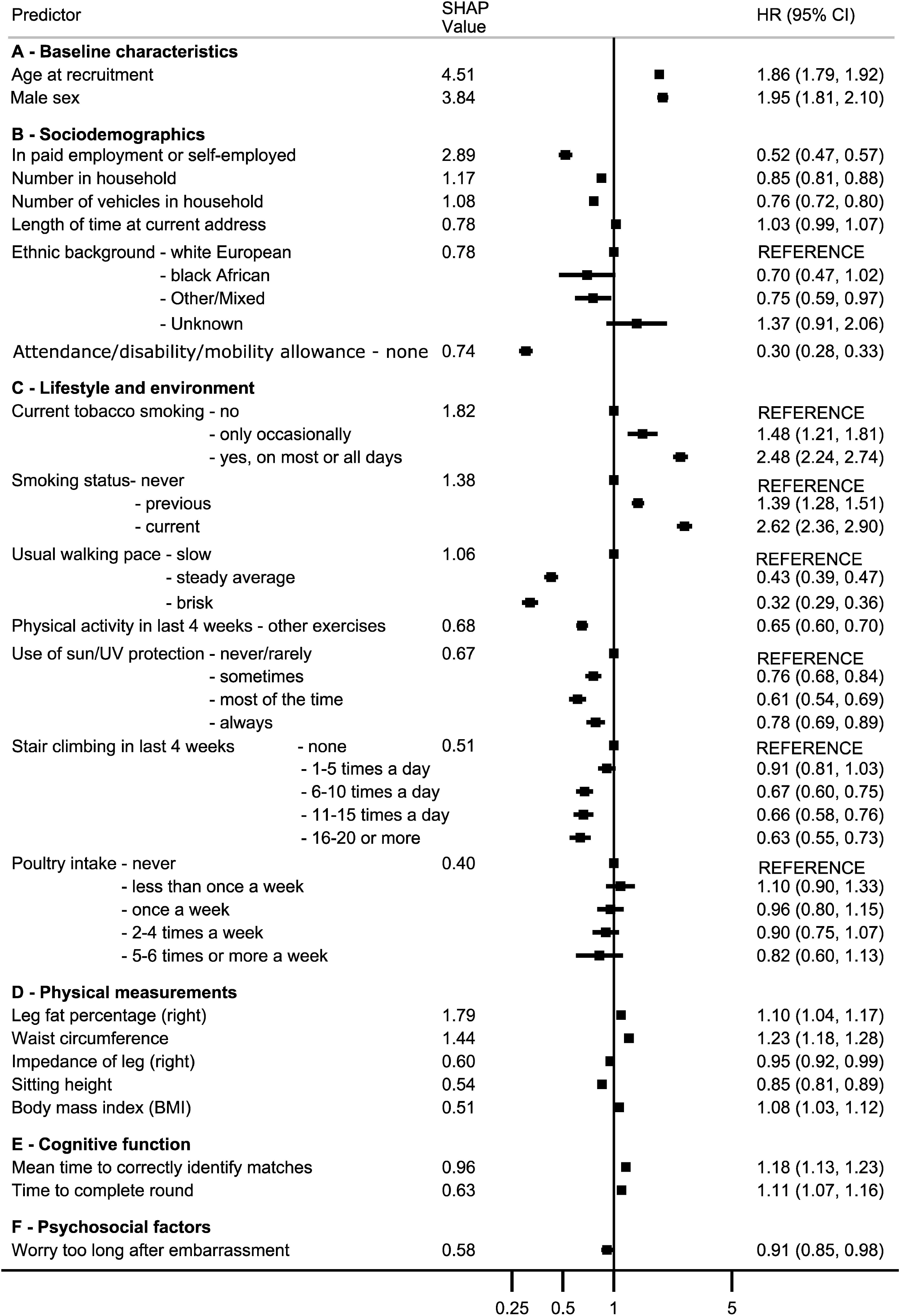
Adjusted Cox regression hazard ratios (HR) with 95% confidence intervals and SHAP values (normalized for 100%) for top 50 predictors ranked by SHAP values belonging to the categories of baseline characteristics, sociodemographics, lifestyle and environment, physical measurements, cognitive functions, and psychosocial factors. Estimates are adjusted for age, sex, Townsend deprivation index, assessment center, and month of birth. The ethnic group “east Asian” is not shown as it had a hazard ratio of 1.4E-20. SHAP: SHapley Additive exPlanation.

**Figure 4.**
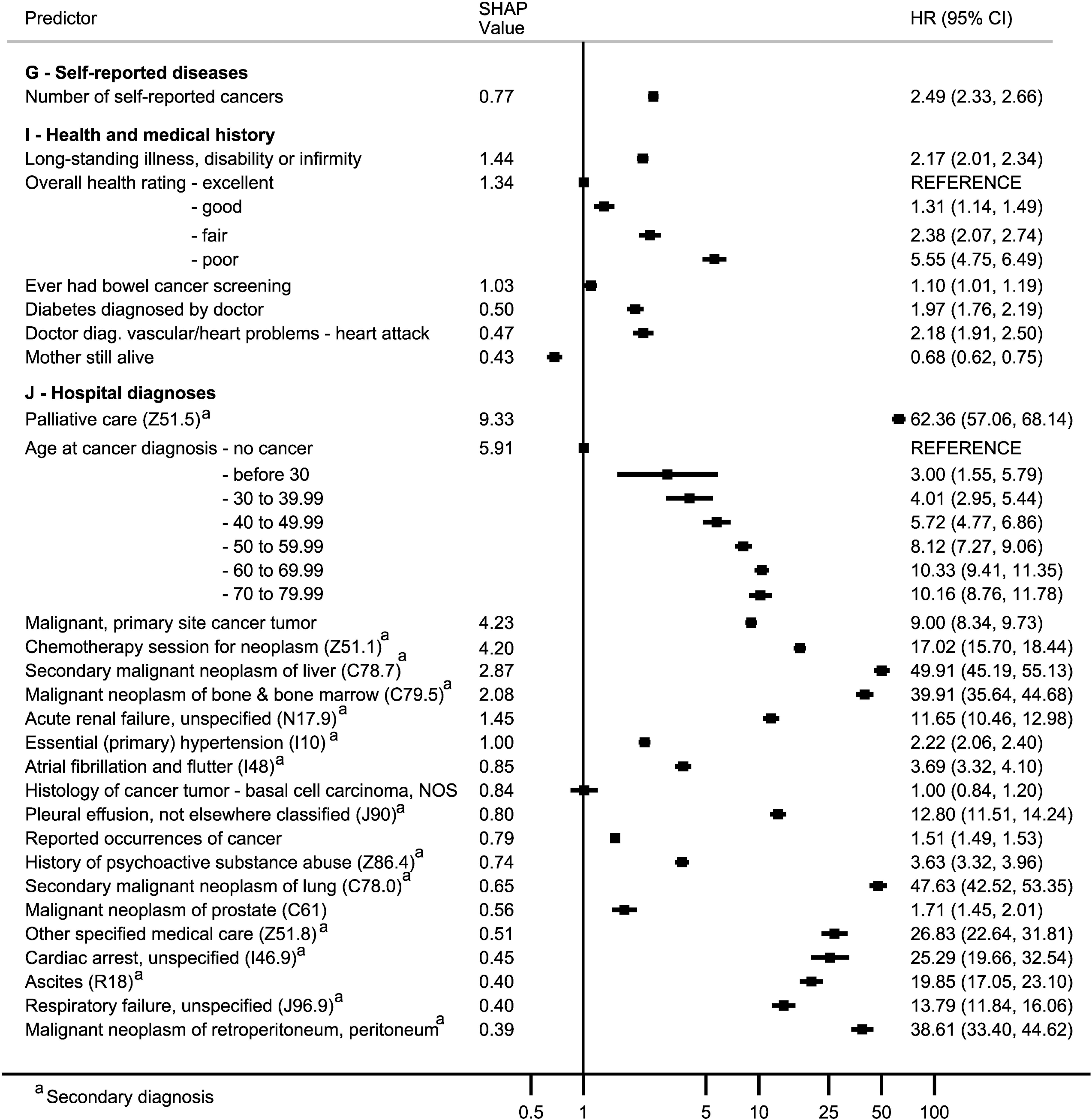
Adjusted Cox regression hazard ratios (HR) with 95% confidence intervals and SHAP values (normalized for 100%) for top 50 predictors ranked by SHAP values belonging to the categories of self-reported diseases, health and medical history and hospital diagnoses. Estimates are adjusted for age, sex, Townsend deprivation index, assessment center, and month of birth. SHAP: SHapley Additive exPlanation.

## DISCUSSION

We examined the value of GBDT-SHAP pipeline in risk factor discovery using mortality prediction in the UK Biobank as the test case. Our test case picked up the expected predictors (e.g. age, sex, palliative care, disease diagnoses) and many other well-known associations (e.g. smoking, social differentials). This demonstrates the effectiveness and viability of GBDT-SHAP pipeline for large-scale hypothesis-free screening in this type of multivariable context where standard epidemiological approaches are not feasible. While our findings here may seem trivial, this approach has particular interest in the context relatively rare diseases for which we know little about, and where large-scale data now provides the first opportunities to identify candidates for prevention.

ML methods are increasingly used in disease prognosis and there is one previous study using ML to predict all-cause mortality in the UK Biobank. This earlier study compared artificial neural network and random forest methods against Cox regression using a set of 60 variables, selected based on their biological plausibility [7]. Where included in our dataset, our hypothesis-free approach, including over 11,000 predictors in the GBDT-SHAP pipeline, picked up all these risk factors or their equivalent. These included key characteristics, such as age, sex, ethnicity, education, Townsend deprivation index, prior cancer diagnoses, smoking, physical activity, blood pressure, diabetes, and adiposity, confirming that our approach is able to identify relevant indicators ‘hidden’ amongst thousands of predictors. While most of the mortality predictors identified in our models were very logical and expected, in a context when less is known about potential predictors, our comprehensive hypothesis-free approach shows great promise for the identification of novel risk factor candidates.

Although GBDT and other ML models tend to be complex and less interpretable than traditional approaches [22, 23], a strength with this approach is the ability to identify relevant risk factors in the context of interactions and non-linear associations. Here, epidemiological analyses require careful model construction which is often impractical when dealing with a very large number of predictors, and complex unknown interactions. Data pre-processing requirements are less for GBDT than that required for standard epidemiological approaches, and our analyses provided examples where non-linear associations which would have remained hidden in standard epidemiological analyses were picked up by the GBDT-SHAP pipeline. Another strength is the ability to incorporate information from thousands of predictors, and to better cope with missing information (without having the need to impute using linear approaches such as MICE[24] or non-linear approaches such as MissForest [25]) in this type of multivariable context. However, in this type of real-life setting, GBDT-SHAP approach is unsuitable for simultaneous inclusion of data from the UKB sub-samples collected after the baseline as participation in the follow-up surveys is correlated with mortality. GBDT-SHAP pipeline also picked up indicators which were associated with the outcome purely due to confounding, as shown by associations of several of the identified ‘important’ predictors being explained by a standard adjustment for baseline factors. Confounding and multicollinearity can also notably affect the SHAP based importance ranking. For example, while the number of cancer diagnoses came among the most important factors, SHAP ranked some cancer diagnoses as less important than age related predictors such as experiencing of knee pain last month, sitting height, and gonarthrosis. For risk factor discovery this may have relatively little importance if at least one relevant indicator is picked up, however, this highlights the importance of replication and more detailed modelling, with caution required when interpreting apparent associations without clear explanations.

Our study demonstrates some of the opportunities in ML based risk factor discovery. There are also limitations, some of which are specific with respect to the dataset. Indeed, reliable analyses from any model require the understanding of the data from which the results are derived. Here, UK Biobank is a cohort of volunteers with higher education and socio-economic status, and lower mortality rates compared to the general population [26]. This type of healthy volunteer bias may affect the external validity of our findings. However, it was reassuring that our data-driven approach identified the traditional risk factors, suggesting the ability to obtain valuable insights in other, less explored settings of risk prediction. Furthermore, as all the analyses in our study were done using a single dataset, we cannot exclude problems with overidentification. Also, as our model used only those predictors available for at least 95% of the participants, we may have left out important determinants which had not been captured in the full cohort. In this observational method exploration, we also cannot establish causal effects, and as we only included adjustments for very basic covariates in our proof of concept test-case, confounding is likely to explain some of the associations.

In conclusion, our data-driven, hypothesis-free approach utilizing ML was a viable, fast, and pragmatic approach to risk factor discovery in a highly phenotyped high dimensional tabular data. Our approach was able to pick up traditional risk factors from among thousands of possible predictors and showed potential for discovering relevant mortality predictors in the context of interactions and non-linear associations. However, to ensure interpretability of the identified predictor – outcome associations, a more detailed modelling utilizing domain expertise and traditional methods is still required.

## Data Availability

Data can be accessed through the UK Biobank

## REFERENCES

1 Obermeyer Z, Emanuel EJ. Predicting the future—big data, machine learning, and clinical medicine. The New England Journal of Medicine 2016;375:1216.

2 Dreiseitl S, Ohno-Machado L. Logistic regression and artificial neural network classification models: a methodology review. Journal of Biomedical Informatics 2002;35:352–9.

3 Weng SF, Reps J, Kai J, et al. Can machine-learning improve cardiovascular risk prediction using routine clinical data? PLOS One 2017;12:e0174944.

4 Hernesniemi JA, Mahdiani S, Tynkkynen JA, et al. Extensive phenotype data and machine learning in prediction of mortality in acute coronary syndrome–the MADDEC study. Annals of Medicine 2019;51:156–63.

5 Blom MC, Ashfaq A, Sant’Anna A, et al. Training machine learning models to predict 30-day mortality in patients discharged from the emergency department: a retrospective, population-based registry study. BMJ Open 2019;9:e028015.

6 Mohamadlou H, Panchavati S, Calvert J, et al. Multicenter validation of a machine-learning algorithm for 48-h all-cause mortality prediction. Health Informatics Journal 2019:1460458219894494.

7 Weng SF, Vaz L, Qureshi N, et al. Prediction of premature all-cause mortality: A prospective general population cohort study comparing machine-learning and standard epidemiological approaches. PLOS One 2019;14:e0214365.

8 Kim SY, Kim S, Cho J, et al. A deep learning model for real-time mortality prediction in critically ill children. Critical Care 2019;23:279.

9 Meyer A, Zverinski D, Pfahringer B, et al. Machine learning for real-time prediction of complications in critical care: a retrospective study. The Lancet Respiratory Medicine 2018;6:905–14.

10 Olson RS, La Cava W, Mustahsan Z, et al. Data-driven advice for applying machine learning to bioinformatics problems. arXiv preprint 170805070 2017.

11 Prokhorenkova L, Gusev G, Vorobev A, et al. CatBoost: unbiased boosting with categorical features. Adv Neur In 2018;31.

12 Lundberg SM, Erion GG, Lee S-I. Consistent individualized feature attribution for tree ensembles. arXiv preprint 180203888 2018.

13 Lundberg SM, Lee S-I. A unified approach to interpreting model predictions. Advances in Neural Information Processing Systems 2017:4765–74.

14 Sudlow C, Gallacher J, Allen N, et al. UK biobank: an open access resource for identifying the causes of a wide range of complex diseases of middle and old age. PLOS Medicine 2015;12:e1001779.

15 Biobank U. UK Biobank Ethics and Governance Framework. UK Biobank 2007.

16 Millard LA, Davies NM, Gaunt TR, et al. Software Application Profile: PHESANT: a tool for performing automated phenome scans in UK Biobank. International Journal of Epidemiology 2017.

17 Ling CX, Sheng VS. Cost-sensitive learning and the class imbalance problem. Encyclopedia of Machine Learning 2008;2011:231–5.

18 Japkowicz N, Stephen S. The class imbalance problem: A systematic study. Intelligent Data Analysis 2002;6:429–49.

19 Maldonado S, Weber R, Famili F. Feature selection for high-dimensional class-imbalanced data sets using support vector machines. Information Sciences 2014;286:228–46.

20 Efron B. Better bootstrap confidence intervals. Journal of the American Statistical Association 1987;82:171–85.

21 Perkins NJ, Schisterman EF. The inconsistency of “optimal” cutpoints obtained using two criteria based on the receiver operating characteristic curve. American Journal of Epidemiology 2006;163:670–5.

22 Beam AL, Kohane IS. Big data and machine learning in health care. JAMA 2018;319:1317–8.

23 Freitas AA. Comprehensible classification models: a position paper. ACM SIGKDD Explorations Newsletter 2014;15:1–10.

24 White IR, Royston P, Wood AM. Multiple imputation using chained equations: issues and guidance for practice. Statistics in Medicine 2011;30:377–99.

25 Stekhoven DJ, Bühlmann P. MissForest—non-parametric missing value imputation for mixed-type data. Bioinformatics 2012;28:112–8.

26 Fry A, Littlejohns TJ, Sudlow C, et al. Comparison of sociodemographic and health-related characteristics of UK Biobank participants with those of the general population. American Journal of Epidemiology 2017;186:1026–34.

