## supplemental materials for "Can we use machine learning to discover risk factors? Testing the proof of principle using data on >11,000 predictors and mortality in the UK Biobank"

**Supplementary Material**

**Contents**

| **Supplementary Methods** | 3 |
| --- | --- |
| **Supplementary Table 1** | 7 |
| **Supplementary Table 2** | 12 |
| **Supplementary Table 3** | 13 |
| **Supplementary Table 4** | 15 |
| **Supplementary Figure 1** | 20 |
| **Supplementary Figure 2** | 21 |
| **Supplementary Figure 3** | 22 |
| **Supplementary Figure 4** | 26 |
| **Supplementary Figure 5** | 31 |

**SUPPLEMENTARY METHODS**

**Background of GBDT**

Gradient boosting decision trees (GBDT) are ensembles of decision trees that are constructed in a sequence such that each subsequent tree after the first tree is trained to predict the error (also called pseudo-residuals) between the observed and predicted value obtained to that point. They are trained using supervised learning. Each decision tree is built by splitting the entire training samples into smaller and smaller groups successively until a predefined condition such as depth of the tree (as determined using a hyper-parameter) or other termination conditions are met. Each training sample will belong to only one leaf of a tree and, in our application, each leaf predicts some level of mortality. Predictors and their values at which the split is to be made to create two new branches to successively build a tree, are chosen by the algorithm in order to optimize an objective function. A development set can be used to avoid overfitting by limiting the number of such decision trees created. GBDTs are currently considered to be the state-of-the-art supervised learning algorithm for building predictive models using tabular data. They have been shown to outperform other machine learning (ML) and traditional statistical methods for mortality predictions in various settings [1-4]. In a more general classification setting, a study conducted to assess the performance of 13 state-of-the-art ML algorithms on a set of 165 publicly available classification problems (mostly bioinformatics problems) found GBDT to be the top ranked in mean ranking across the problems [5]. There are end-to-end implementations of GBDTs, capable of handling billions of samples and millions of variables such as XGBoost, LightGBM, and CatBoost. They utilize graphical processing units (GPUs) in addition to central processing units to improve training and inference speed.

Each ML algorithm has its own hyperparameters, also known as tuning parameters and that can control the behavior of an algorithm. Examples of such hyperparameters include learning rate, depth of decision trees, maximum number of decision trees to be created and so on. Generally, CatBoost [6] outperforms XGBoost and LightGBM in performance using default hyperparameters [7, 8]. Also, improvements in performance using tuned hyperparameters may not be largely different from performance with default hyperparameters as seen in some studies using CatBoost [8]. CatBoost provides native support to handle categorical predictors (with or without numeric values). It can be instructed to consider missing value as an instance of value, guaranteeing a split between missing value and other non-missing values while trees are built.

**Specific GBDT methods**

For CatBoost, the most important hyperparameters are learning rate and number of trees to be built (also known as number of iterations or number of estimators). If used with default values, CatBoost dynamically selects learning rate based on number of iterations (default value of iterations is 1,000). For our experiments, we set the number of iterations to 10,000 and used a development set to avoid overfitting by stopping growing new trees when there is no improvement in area under the receiver operating characteristics curve (AUROC) performance on the development set in 50 consecutive iterations. Models were set to utilize GPUs and boosting type was set to plain boosting. All other hyper-parameters were left at default values. We used predictors without imputing missing values as we used only baseline predictors (which were available for at least 95% of the participants) and hospital diagnoses.

**Pre-processing using PHESANT**

PHESANT (PHEnome Scan Analysis) [9] classifies variables as continuous, ordinal, and categorical using a rule-based system to determine the appropriate coding of each variable. It also deals with various scenarios such as handling of multiple initial measurement of variables (e.g. spirometry), coding unusual values (e.g. negative values are used to code answers such as ‘Preferred not to answer’ and ‘Don’t know’) as missing, changing the order of variable values to make them logical (e.g., field 1239, current smoking status) and creating proper dummy variables (e.g., secondary diagnoses field 41204 had 184 array elements and any ICD10 code could be stored in any array elements).

**SHAP (SHapley Additive exPlanation) Values**

SHAP values are based on Shapley values (derived by Lloyd Shapley in 1953 [10]), a solution concept in game theory. Shapley values deal with how fairly (by satisfying certain conditions) the payoffs from a cooperative game can be distributed to game players. Shapley value for a player is defined as the average marginal contribution of that player, considering all possible combinations that the player can be part of.

SHAP values is a local additive feature (features and predictors are used here synonymously) attribution method [11], i.e., it contrastively explains each observation in isolation using a linear function with predicted output as function output and simplified and interpretable feature values as input. Since SHAP values use a linear approximation, for binary classifiers transforming margins using logistic function, SHAP values will be in log-odds space. SHAP values are consistent with respect to feature attribution measurement in tree models, even in the presence of correlated features. If *f* represents the ML model learned (e.g., a trained GBDT model) and *g* represents the local linear explanation model (for a particular observation, $\boldsymbol{x}$), then

$f\left( \boldsymbol{x} \right)=g\left( \boldsymbol{x}^{'} \right)= \emptyset_{0}+ \sum_{i=1}^{M} \emptyset_{i}x_{i}^{'}$,

where, $\emptyset_{0}$ is the output when no input is present, *M* is the number of features, $\boldsymbol{x}^{\boldsymbol{'}}$ is the simplified, dichotomized and interpretable vector representing ***x*** in the local explanation model and $\emptyset_{i}\mathbb{\in R}$ is the attribution to each feature. According to [11], $\emptyset_{i}$ given *f* and ***x*** and satisfying some desirable conditions (local consistency and missingness) is given as [11]

$$\emptyset_{i}\left( f,\boldsymbol{x} \right)= \sum_{\boldsymbol{z}^{\boldsymbol{'}}\subseteq\boldsymbol{x}^{'}} \frac{\left| \boldsymbol{z}^{'} \right|!\left( M-\left| \boldsymbol{z}^{'} \right|-1 \right)!}{M!} \left[ f_{\boldsymbol{x}}\left( \boldsymbol{z}^{'} \right)- f_{\boldsymbol{x}}\left( \boldsymbol{z}^{'}\backslash i \right) \right],$$

where, $\left| \boldsymbol{z}^{'} \right|$ is the number of features present (non-zero elements) in $\boldsymbol{x}^{\boldsymbol{'}}$. The solution to the above equation is the Shapley values of a conditional expectation function of the original model *f*. We use the recent implementation, called TreeSHAP [12] specifically developed for tree-based models such as random forest and gradient boosting. TreeSHAP can calculate SHAP values much faster by reducing computational complexity from $\mathcal{O}\left( TL2^{M} \right)$ to $\mathcal{O}\left( TLD^{2} \right)$ (*T* is the number of trees, *L* is the number of leaves, *M* is the number of features, and *D* is the depth of the tree) compared to the previous implementation KernalSHAP [11]. We used the Python package ‘SHAP’ version 0.34 for calculating SHAP values.

**Variable selection using SHAP values and correlation**

For each predictor, we calculated variable importance as the mean absolute SHAP value in the training set, as

$\varphi_{i}= \frac{1}{N} \sum_{j=1}^{N} |\emptyset_{ji}|$,

where *N* is the total number of observations in the training set.

We normalized mean absolute SHAP values ($\varphi_{i})$ to 100% and a cut-off value of 0.05% was used to identify ‘important’ predictors. We used Spearman’s ρ (above 0.9) to identify sets of highly correlated predictors and removed all but one (the one recorded for the greatest number of samples) from those sets to produce the final set of important predictors. We plotted SHAP values of important predictors for each sample to understand the direction and magnitude of impact of individual predictors on model output.

### Supplementary Table 1. List of UK Biobank baseline and hospital diagnoses fields considered in the study. There were in total 177 UK Biobank fields (without considering array elements within a field to store multiple values. For example, the field 42104 for recording secondary diagnoses using ICD10 codes had 184 array elements). The baseline fields selected were available for at least 95% of the participants.

| **UK biobank field ID** | **Field description** | **Hierarchical category ^a^** |
| --- | --- | --- |
| **A - Baseline characteristics** | |  |
| 31 | Sex | Population characteristics > Baseline characteristics |
| 34 | Year of birth | Population characteristics > Baseline characteristics |
| 52 | Month of birth | Population characteristics > Baseline characteristics |
| 189 | Townsend deprivation index at recruitment | Population characteristics > Baseline characteristics |
| 21003 | Age when attended assessment center | UK Biobank Assessment Centre > Recruitment > Reception |
| 21022 | Age at recruitment | Population characteristics > Baseline characteristics |
| **B - Sociodemographics** | |  |
| 670 | Type of accommodation lived in | UK Biobank Assessment Centre > Touchscreen > Sociodemographics > Household |
| 680 | Own or rent accommodation lived in | UK Biobank Assessment Centre > Touchscreen > Sociodemographics > Household |
| 699 | Length of time at current address | UK Biobank Assessment Centre > Touchscreen > Sociodemographics > Household |
| 709 | Number in household | UK Biobank Assessment Centre > Touchscreen > Sociodemographics > Household |
| 728 | Number of vehicles in household | UK Biobank Assessment Centre > Touchscreen > Sociodemographics > Household |
| 6138 | Qualifications | UK Biobank Assessment Centre > Touchscreen > Sociodemographics > Education |
| 6139 | Gas or solid-fuel cooking/heating | UK Biobank Assessment Centre > Touchscreen > Sociodemographics > Household |
| 6142 | Current employment status | UK Biobank Assessment Centre > Touchscreen > Sociodemographics > Employment |
| 6146 | Attendance/disability/mobility allowance | UK Biobank Assessment Centre > Touchscreen > Sociodemographics > Other sociodemographic factors |
| 21000 | Ethnic background | UK Biobank Assessment Centre > Touchscreen > Sociodemographics > Ethnicity |
| **C - Lifestyle and environment** | |  |
| 120 | Birth weight known | UK Biobank Assessment Centre > Verbal interview > Early life factors |
| 864 | Number of days/week walked 10+ minutes | UK Biobank Assessment Centre > Touchscreen > Lifestyle and environment > Physical activity |
| 924 | Usual walking pace | UK Biobank Assessment Centre > Touchscreen > Lifestyle and environment > Physical activity |
| 943 | Frequency of stair climbing in last 4 weeks | UK Biobank Assessment Centre > Touchscreen > Lifestyle and environment > Physical activity |
| 1239 | Current tobacco smoking | UK Biobank Assessment Centre > Touchscreen > Lifestyle and environment > Smoking |
| 1289 | Cooked vegetable intake | UK Biobank Assessment Centre > Touchscreen > Lifestyle and environment > Diet |
| 1309 | Fresh fruit intake | UK Biobank Assessment Centre > Touchscreen > Lifestyle and environment > Diet |
| 1329 | Oily fish intake | UK Biobank Assessment Centre > Touchscreen > Lifestyle and environment > Diet |
| 1339 | Non-oily fish intake | UK Biobank Assessment Centre > Touchscreen > Lifestyle and environment > Diet |
| 1349 | Processed meat intake | UK Biobank Assessment Centre > Touchscreen > Lifestyle and environment > Diet |
| 1359 | Poultry intake | UK Biobank Assessment Centre > Touchscreen > Lifestyle and environment > Diet |
| 1369 | Beef intake | UK Biobank Assessment Centre > Touchscreen > Lifestyle and environment > Diet |
| 1379 | Lamb/mutton intake | UK Biobank Assessment Centre > Touchscreen > Lifestyle and environment > Diet |
| 1389 | Pork intake | UK Biobank Assessment Centre > Touchscreen > Lifestyle and environment > Diet |
| 1408 | Cheese intake | UK Biobank Assessment Centre > Touchscreen > Lifestyle and environment > Diet |
| 1418 | Milk type used | UK Biobank Assessment Centre > Touchscreen > Lifestyle and environment > Diet |
| 1428 | Spread type | UK Biobank Assessment Centre > Touchscreen > Lifestyle and environment > Diet |
| 1438 | Bread intake | UK Biobank Assessment Centre > Touchscreen > Lifestyle and environment > Diet |
| 1448 | Bread type | UK Biobank Assessment Centre > Touchscreen > Lifestyle and environment > Diet |
| 1478 | Salt added to food | UK Biobank Assessment Centre > Touchscreen > Lifestyle and environment > Diet |
| 1488 | Tea intake | UK Biobank Assessment Centre > Touchscreen > Lifestyle and environment > Diet |
| 1518 | Hot drink temperature | UK Biobank Assessment Centre > Touchscreen > Lifestyle and environment > Diet |
| 1538 | Major dietary changes in the last 5 years | UK Biobank Assessment Centre > Touchscreen > Lifestyle and environment > Diet |
| 1548 | Variation in diet | UK Biobank Assessment Centre > Touchscreen > Lifestyle and environment > Diet |
| 1647 | Country of birth (UK/elsewhere) | UK Biobank Assessment Centre > Touchscreen > Early life factors |
| 1687 | Comparative body size at age 10 | UK Biobank Assessment Centre > Touchscreen > Early life factors |
| 1697 | Comparative height size at age 10 | UK Biobank Assessment Centre > Touchscreen > Early life factors |
| 1707 | Handedness (chirality/laterality) | UK Biobank Assessment Centre > Touchscreen > Early life factors |
| 1717 | Skin color | UK Biobank Assessment Centre > Touchscreen > Lifestyle and environment > Sun exposure |
| 1727 | Ease of skin tanning | UK Biobank Assessment Centre > Touchscreen > Lifestyle and environment > Sun exposure |
| 1747 | Hair color (natural, before greying) | UK Biobank Assessment Centre > Touchscreen > Lifestyle and environment > Sun exposure |
| 1767 | Adopted as a child | UK Biobank Assessment Centre > Touchscreen > Early life factors |
| 1777 | Part of a multiple birth | UK Biobank Assessment Centre > Touchscreen > Early life factors |
| 2267 | Use of sun/UV protection | UK Biobank Assessment Centre > Touchscreen > Lifestyle and environment > Sun exposure |
| 6144 | Never eat eggs, dairy, wheat, sugar | UK Biobank Assessment Centre > Touchscreen > Lifestyle and environment > Diet |
| 6162 | Types of transport used (excluding work) | UK Biobank Assessment Centre > Touchscreen > Lifestyle and environment > Physical activity |
| 6164 | Types of physical activity in last 4 weeks | UK Biobank Assessment Centre > Touchscreen > Lifestyle and environment > Physical activity |
| 20116 | Smoking status | UK Biobank Assessment Centre > Touchscreen > Lifestyle and environment > Smoking |
| 20160 | Ever smoked | UK Biobank Assessment Centre > Touchscreen > Lifestyle and environment > Smoking |
| **D - Physical measurements** | |  |
| 21 | Weight method | UK Biobank Assessment Centre > Physical measures > Anthropometry > Body size measures |
| 46 | Hand grip strength (left) | UK Biobank Assessment Centre > Physical measures > Hand grip strength |
| 47 | Hand grip strength (right) | UK Biobank Assessment Centre > Physical measures > Hand grip strength |
| 48 | Waist circumference | UK Biobank Assessment Centre > Physical measures > Anthropometry > Body size measures |
| 49 | Hip circumference | UK Biobank Assessment Centre > Physical measures > Anthropometry > Body size measures |
| 50 | Standing height | UK Biobank Assessment Centre > Physical measures > Anthropometry > Body size measures |
| 3088 | Contra-indications for spirometry | UK Biobank Assessment Centre > Physical measures > Spirometry |
| 20015 | Sitting height | UK Biobank Assessment Centre > Physical measures > Anthropometry > Body size measures |
| 21001 | Body mass index (BMI) | UK Biobank Assessment Centre > Physical measures > Anthropometry > Body size measures |
| 21002 | Weight | UK Biobank Assessment Centre > Physical measures > Anthropometry > Body size measures |
| 23098 | Weight | UK Biobank Assessment Centre > Physical measures > Anthropometry > Impedance measures |
| 23099 | Body fat percentage | UK Biobank Assessment Centre > Physical measures > Anthropometry > Impedance measures |
| 23100 | Whole body fat mass | UK Biobank Assessment Centre > Physical measures > Anthropometry > Impedance measures |
| 23101 | Whole body fat-free mass | UK Biobank Assessment Centre > Physical measures > Anthropometry > Impedance measures |
| 23102 | Whole body water mass | UK Biobank Assessment Centre > Physical measures > Anthropometry > Impedance measures |
| 23104 | Body mass index (BMI) | UK Biobank Assessment Centre > Physical measures > Anthropometry > Impedance measures |
| 23105 | Basal metabolic rate | UK Biobank Assessment Centre > Physical measures > Anthropometry > Impedance measures |
| 23106 | Impedance of whole body | UK Biobank Assessment Centre > Physical measures > Anthropometry > Impedance measures |
| 23107 | Impedance of leg (right) | UK Biobank Assessment Centre > Physical measures > Anthropometry > Impedance measures |
| 23108 | Impedance of leg (left) | UK Biobank Assessment Centre > Physical measures > Anthropometry > Impedance measures |
| 23109 | Impedance of arm (right) | UK Biobank Assessment Centre > Physical measures > Anthropometry > Impedance measures |
| 23110 | Impedance of arm (left) | UK Biobank Assessment Centre > Physical measures > Anthropometry > Impedance measures |
| 23111 | Leg fat percentage (right) | UK Biobank Assessment Centre > Physical measures > Anthropometry > Impedance measures |
| 23112 | Leg fat mass (right) | UK Biobank Assessment Centre > Physical measures > Anthropometry > Impedance measures |
| 23113 | Leg fat-free mass (right) | UK Biobank Assessment Centre > Physical measures > Anthropometry > Impedance measures |
| 23114 | Leg predicted mass (right) | UK Biobank Assessment Centre > Physical measures > Anthropometry > Impedance measures |
| 23115 | Leg fat percentage (left) | UK Biobank Assessment Centre > Physical measures > Anthropometry > Impedance measures |
| 23116 | Leg fat mass (left) | UK Biobank Assessment Centre > Physical measures > Anthropometry > Impedance measures |
| 23117 | Leg fat-free mass (left) | UK Biobank Assessment Centre > Physical measures > Anthropometry > Impedance measures |
| 23118 | Leg predicted mass (left) | UK Biobank Assessment Centre > Physical measures > Anthropometry > Impedance measures |
| 23119 | Arm fat percentage (right) | UK Biobank Assessment Centre > Physical measures > Anthropometry > Impedance measures |
| 23120 | Arm fat mass (right) | UK Biobank Assessment Centre > Physical measures > Anthropometry > Impedance measures |
| 23121 | Arm fat-free mass (right) | UK Biobank Assessment Centre > Physical measures > Anthropometry > Impedance measures |
| 23122 | Arm predicted mass (right) | UK Biobank Assessment Centre > Physical measures > Anthropometry > Impedance measures |
| 23123 | Arm fat percentage (left) | UK Biobank Assessment Centre > Physical measures > Anthropometry > Impedance measures |
| 23124 | Arm fat mass (left) | UK Biobank Assessment Centre > Physical measures > Anthropometry > Impedance measures |
| 23125 | Arm fat-free mass (left) | UK Biobank Assessment Centre > Physical measures > Anthropometry > Impedance measures |
| 23126 | Arm predicted mass (left) | UK Biobank Assessment Centre > Physical measures > Anthropometry > Impedance measures |
| 23127 | Trunk fat percentage | UK Biobank Assessment Centre > Physical measures > Anthropometry > Impedance measures |
| 23128 | Trunk fat mass | UK Biobank Assessment Centre > Physical measures > Anthropometry > Impedance measures |
| 23129 | Trunk fat-free mass | UK Biobank Assessment Centre > Physical measures > Anthropometry > Impedance measures |
| 23130 | Trunk predicted mass | UK Biobank Assessment Centre > Physical measures > Anthropometry > Impedance measures |
| **E - Cognitive function** | |  |
| 398 | Number of correct matches in round | UK Biobank Assessment Centre > Cognitive function > Pairs matching |
| 399 | Number of incorrect matches in round | UK Biobank Assessment Centre > Cognitive function > Pairs matching |
| 400 | Time to complete round | UK Biobank Assessment Centre > Cognitive function > Pairs matching |
| 403 | Number of times snap-button pressed | UK Biobank Assessment Centre > Cognitive function > Reaction time |
| 404 | Duration to first press of snap-button in each round | UK Biobank Assessment Centre > Cognitive function > Reaction time |
| 20023 | Mean time to correctly identify matches | UK Biobank Assessment Centre > Cognitive function > Reaction time |
| **F - Psychosocial factors** | |  |
| 1920 | Mood swings | UK Biobank Assessment Centre > Touchscreen > Psychosocial factors > Mental health |
| 1930 | Miserableness | UK Biobank Assessment Centre > Touchscreen > Psychosocial factors > Mental health |
| 1940 | Irritability | UK Biobank Assessment Centre > Touchscreen > Psychosocial factors > Mental health |
| 1950 | Sensitivity / hurt feelings | UK Biobank Assessment Centre > Touchscreen > Psychosocial factors > Mental health |
| 1960 | Fed-up feelings | UK Biobank Assessment Centre > Touchscreen > Psychosocial factors > Mental health |
| 1970 | Nervous feelings | UK Biobank Assessment Centre > Touchscreen > Psychosocial factors > Mental health |
| 1980 | Worrier / anxious feelings | UK Biobank Assessment Centre > Touchscreen > Psychosocial factors > Mental health |
| 1990 | Tense / 'highly strung' | UK Biobank Assessment Centre > Touchscreen > Psychosocial factors > Mental health |
| 2000 | Worry too long after embarrassment | UK Biobank Assessment Centre > Touchscreen > Psychosocial factors > Mental health |
| 2010 | Suffer from 'nerves' | UK Biobank Assessment Centre > Touchscreen > Psychosocial factors > Mental health |
| 2020 | Loneliness, isolation | UK Biobank Assessment Centre > Touchscreen > Psychosocial factors > Mental health |
| 2030 | Guilty feelings | UK Biobank Assessment Centre > Touchscreen > Psychosocial factors > Mental health |
| 2040 | Risk taking | UK Biobank Assessment Centre > Touchscreen > Psychosocial factors > Mental health |
| 2050 | Frequency of depressed mood in last 2 weeks | UK Biobank Assessment Centre > Touchscreen > Psychosocial factors > Mental health |
| 2060 | Frequency of unenthusiasm / disinterest in last 2 weeks | UK Biobank Assessment Centre > Touchscreen > Psychosocial factors > Mental health |
| 2070 | Frequency of tenseness / restlessness in last 2 weeks | UK Biobank Assessment Centre > Touchscreen > Psychosocial factors > Mental health |
| 2080 | Frequency of tiredness / lethargy in last 2 weeks | UK Biobank Assessment Centre > Touchscreen > Psychosocial factors > Mental health |
| 2090 | Seen doctor (GP) for nerves, anxiety, tension, or depression | UK Biobank Assessment Centre > Touchscreen > Psychosocial factors > Mental health |
| 2100 | Seen a psychiatrist for nerves, anxiety, tension, or depression | UK Biobank Assessment Centre > Touchscreen > Psychosocial factors > Mental health |
| 6145 | Illness, injury, bereavement, stress in last 2 years | UK Biobank Assessment Centre > Touchscreen > Psychosocial factors > Mental health |
| **G - Self-reported diseases** | |  |
| 134 | Number of self-reported cancers | UK Biobank Assessment Centre > Verbal interview > Medical conditions |
| 135 | Number of self-reported non-cancer illnesses | UK Biobank Assessment Centre > Verbal interview > Medical conditions |
| **H - Medications & Operations** | |  |
| 136 | Number of operations, self-reported | UK Biobank Assessment Centre > Verbal interview > Operations |
| 137 | Number of treatments/medications taken | UK Biobank Assessment Centre > Verbal interview > Medications |
| 3079 | Pacemaker | UK Biobank Assessment Centre > Verbal interview > Operations |
| **I - Health and medical history** | |  |
| 1797 | Father still alive | UK Biobank Assessment Centre > Touchscreen > Family history |
| 1835 | Mother still alive | UK Biobank Assessment Centre > Touchscreen > Family history |
| 1873 | Number of full brothers | UK Biobank Assessment Centre > Touchscreen > Family history |
| 1883 | Number of full sisters | UK Biobank Assessment Centre > Touchscreen > Family history |
| 2178 | Overall health rating | UK Biobank Assessment Centre > Touchscreen > Health and medical history > General health |
| 2188 | Long-standing illness, disability, or infirmity | UK Biobank Assessment Centre > Touchscreen > Health and medical history > General health |
| 2207 | Wears glasses or contact lenses | UK Biobank Assessment Centre > Touchscreen > Health and medical history > Eyesight |
| 2227 | Other eye problems | UK Biobank Assessment Centre > Touchscreen > Health and medical history > Eyesight |
| 2247 | Hearing difficulty/problems | UK Biobank Assessment Centre > Touchscreen > Health and medical history > Hearing |
| 2257 | Hearing difficulty/problems with background noise | UK Biobank Assessment Centre > Touchscreen > Health and medical history > Hearing |
| 2296 | Falls in the last year | UK Biobank Assessment Centre > Touchscreen > Health and medical history > General health |
| 2306 | Weight change compared with 1 year ago | UK Biobank Assessment Centre > Touchscreen > Health and medical history > General health |
| 2316 | Wheeze or whistling in the chest in last year | UK Biobank Assessment Centre > Touchscreen > Health and medical history > Breathing |
| 2335 | Chest pain or discomfort | UK Biobank Assessment Centre > Touchscreen > Health and medical history > Chest pain |
| 2345 | Ever had bowel cancer screening | UK Biobank Assessment Centre > Touchscreen > Health and medical history > Cancer screening |
| 2443 | Diabetes diagnosed by doctor | UK Biobank Assessment Centre > Touchscreen > Health and medical history > Medical conditions |
| 2453 | Cancer diagnosed by doctor | UK Biobank Assessment Centre > Touchscreen > Health and medical history > Medical conditions |
| 2463 | Fractured/broken bones in last 5 years | UK Biobank Assessment Centre > Touchscreen > Health and medical history > Medical conditions |
| 2473 | Other serious medical condition/disability diagnosed by doctor | UK Biobank Assessment Centre > Touchscreen > Health and medical history > Medical conditions |
| 2492 | Taking other prescription medications | UK Biobank Assessment Centre > Touchscreen > Health and medical history > Medication |
| 6149 | Mouth/teeth dental problems | UK Biobank Assessment Centre > Touchscreen > Health and medical history > Mouth |
| 6150 | Vascular/heart problems diagnosed by doctor | UK Biobank Assessment Centre > Touchscreen > Health and medical history > Medical conditions |
| 6152 | Blood clot, DVT, bronchitis, emphysema, asthma, rhinitis, eczema, allergy diagnosed by doctor | UK Biobank Assessment Centre > Touchscreen > Health and medical history > Medical conditions |
| 6154 | Medication for pain relief, constipation, heartburn | UK Biobank Assessment Centre > Touchscreen > Health and medical history > Medication |
| 6155 | Vitamin and mineral supplements | UK Biobank Assessment Centre > Touchscreen > Health and medical history > Medication |
| 6159 | Pain type(s) experienced in last month | UK Biobank Assessment Centre > Touchscreen > Health and medical history > Pain |
| 6179 | Mineral and other dietary supplements | UK Biobank Assessment Centre > Touchscreen > Health and medical history > Medication |
| **J - Hospital diagnoses** | |  |
| 40006 | Type of cancer: ICD10 | Health-related outcomes > Cancer register |
| 40008 | Age at cancer diagnosis | Health-related outcomes > Cancer register |
| 40009 | Reported occurrences of cancer | Health-related outcomes > Cancer register |
| 40011 | Histology of cancer tumor | Health-related outcomes > Cancer register |
| 40012 | Behavior of cancer tumor | Health-related outcomes > Cancer register |
| 40013 | Type of cancer: ICD9 | Health-related outcomes > Cancer register |
| 40019 | Cancer report format | Health-related outcomes > Cancer register |
| 41201 | External causes - ICD10 | Health-related outcomes > Hospital inpatient > Summary Diagnoses |
| 41202 | Diagnoses - main ICD10 | Health-related outcomes > Hospital inpatient > Summary Diagnoses |
| 41203 | Diagnoses - main ICD9 | Health-related outcomes > Hospital inpatient > Summary Diagnoses |
| 41204 | Diagnoses - secondary ICD10 | Health-related outcomes > Hospital inpatient > Summary Diagnoses |
| 41205 | Diagnoses - secondary ICD9 | Health-related outcomes > Hospital inpatient > Summary Diagnoses |
| 41219 | Anesthetics administered during delivery | Health-related outcomes > Hospital inpatient > Summary Maternity |
| 41220 | Anesthetics administered post delivery | Health-related outcomes > Hospital inpatient > Summary Maternity |
| 41221 | Delivery methods | Health-related outcomes > Hospital inpatient > Summary Maternity |
| 41222 | Delivery onset methods | Health-related outcomes > Hospital inpatient > Summary Maternity |
| 41223 | Delivery places | Health-related outcomes > Hospital inpatient > Summary Maternity |
| 41224 | Intended delivery places | Health-related outcomes > Hospital inpatient > Summary Maternity |
| 41225 | Resuscitation methods | Health-related outcomes > Hospital inpatient > Summary Maternity |
| 41226 | Sex of baby | Health-related outcomes > Hospital inpatient > Summary Maternity |
| 41227 | Status of baby at birth | Health-related outcomes > Hospital inpatient > Summary Maternity |
| 41228 | Statuses of person conducting delivery | Health-related outcomes > Hospital inpatient > Summary Maternity |

^a^ More information on UK biobank field categories can be found at the UK biobank website (<https://biobank.ndph.ox.ac.uk/showcase/cats.cgi>)

**Supplementary Table 2.** Category wise count of predictors before and after pre-processing using PHESANT (PHEnome Scan Analysis) software package. Important predictors are the predictors identified using SHAP (SHapley Additive exPlanation) values passing the selected SHAP value threshold of 0.05% and after further elimination of predictors using Spearman’s ρ.

| **Category** | **All predictors** ^a^ | | **Important predictors** ^b^ | |
| --- | --- | --- | --- | --- |
|  | **UK Biobank fields** | **Derived predictors** ^c^ | **UK Biobank fields** | **Derived predictors** ^c^ |
| A - Baseline characteristics | 6 | 6 | 4 | 4 |
| B - Sociodemographics | 10 | 29 | 9 | 15 |
| C - Lifestyle and environment | 39 | 51 | 25 | 28 |
| D - Physical measurements | 42 | 42 | 13 | 13 |
| E - Cognitive function | 6 | 6 | 3 | 3 |
| F - Psychosocial factors | 20 | 26 | 8 | 10 |
| G - Self-reported diseases | 2 | 2 | 2 | 2 |
| H - Medications & Operations | 3 | 3 | 2 | 2 |
| I - Health and medical history | 27 | 69 | 21 | 31 |
| J - Hospital diagnoses | 22 | 11,405 | 8 | 85 |
| **Total** | **177** | **11,639** | **95** | **193** |

^a^ Fields considered in the gradient boosting decision tree (GBDT) models with all predictors in its input.

^b^ Fields considered in the GBDT models with only important predictors in its input.

^c^ Derived predictors are the output predictors of PHESANT pre-processing and were used as input to GBDT models.

**Supplementary Table 3.** Category wise listing of important predictors identified. Three sets of SHAP values are provided: a) SHAP values (normalized for 100%) from a gradient boosting decision trees (GBDT) model with all predictors in its input totaling 84%, b) SHAP values adjusted for a total of 100% and c) SHAP values from a GBDT model with only important predictors in its input.

| **Predictor** ^a^ | **SHAP value**  **(all)**^b^ | **SHAP value**  **(all adj)** ^c^ | **SHAP value**  **(im.)** ^d^ |
| --- | --- | --- | --- |
| **A - Baseline characteristics** |  |  |  |
| Age at recruitment | 0.46 | 0.55 | 4.51 |
| Sex—male | 3.02 | 3.59 | 3.84 |
| Townsend deprivation index | 0.20 | 0.24 | 0.26 |
| Month of birth | 0.30 | 0.36 | 0.23 |
| **A - Baseline characteristics Total** | **3.98** | **4.74** | **8.85** |
| **B - Sociodemographics** |  |  |  |
| In paid employment or self-employed | 2.64 | 3.15 | 2.89 |
| Number in household | 1.17 | 1.39 | 1.17 |
| Number of vehicles in household | 1.15 | 1.37 | 1.08 |
| Length of time at current address | 0.67 | 0.80 | 0.78 |
| Ethnic background | 0.57 | 0.68 | 0.78 |
| Attendance/disability/mobility allowance—none | 0.75 | 0.89 | 0.74 |
| Attendance/disability/mobility allowance - disability | 0.25 | 0.29 | 0.21 |
| Gas or solid-fuel cooking/heating - a gas fire that you use regularly in winter time | 0.14 | 0.17 | 0.19 |
| Gas or solid-fuel cooking/heating - an open solid fuel fire that you use regularly in winter time | 0.10 | 0.12 | 0.19 |
| Current employment status - unable to work because of sickness or disability | 0.24 | 0.29 | 0.18 |
| Qualifications - CSEs or equivalent | 0.08 | 0.09 | 0.16 |
| Qualifications - A levels/AS levels or equivalent | 0.07 | 0.09 | 0.13 |
| Type of accommodation lived in—house, flat, mobile, sheltered, care home | 0.05 | 0.06 | 0.13 |
| Gas or solid-fuel cooking/heating—none | 0.08 | 0.09 | 0.12 |
| Current employment status - unemployed | 0.07 | 0.08 | 0.03 |
| **B - Sociodemographics Total** | **8.03** | **9.56** | **8.80** |
| **C - Lifestyle and environment** |  |  |  |
| Current tobacco smoking—more frequently | 1.80 | 2.14 | 1.82 |
| Smoking status—never, previous, current | 1.41 | 1.67 | 1.38 |
| Usual walking pace | 1.06 | 1.27 | 1.06 |
| Physical activity in last 4 weeks - other exercises (e.g.: swimming, cycling, keep fit, bowling) | 0.78 | 0.92 | 0.68 |
| Use of sun/UV protection—always | 0.62 | 0.74 | 0.67 |
| Frequency of stair climbing in last 4 weeks | 0.45 | 0.54 | 0.51 |
| Poultry intake | 0.30 | 0.36 | 0.40 |
| Tea intake | 0.35 | 0.42 | 0.39 |
| Processed meat intake | 0.27 | 0.32 | 0.33 |
| Ever smoked | 0.24 | 0.28 | 0.29 |
| Beef intake | 0.25 | 0.30 | 0.26 |
| Light DIY physical activity in last 4 weeks | 0.20 | 0.24 | 0.24 |
| Cooked vegetable intake | 0.27 | 0.32 | 0.22 |
| Bread intake | 0.11 | 0.13 | 0.22 |
| Eat eggs, diary, wheat, sugar2 | 0.12 | 0.14 | 0.21 |
| Bread type | 0.19 | 0.23 | 0.21 |
| Comparative height size at age 10 | 0.16 | 0.19 | 0.21 |
| Types of physical activity in last 4 weeks - Heavy DIY (e.g.: weeding, lawn mowing, carpentry, digging) | 0.16 | 0.19 | 0.20 |
| Pork intake | 0.14 | 0.16 | 0.19 |
| Number of days/week walked 10+ minutes | 0.13 | 0.15 | 0.17 |
| Skin color—less fair | 0.11 | 0.14 | 0.16 |
| Types of physical activity in last 4 weeks - strenuous sports | 0.05 | 0.06 | 0.15 |
| Salt added to food | 0.10 | 0.12 | 0.15 |
| Cheese intake | 0.07 | 0.09 | 0.14 |
| Country of birth (UK/elsewhere) | 0.13 | 0.16 | 0.13 |
| Handedness—right-handed, left-handed, use both hands | 0.07 | 0.09 | 0.12 |
| Hot drink temperature—less hot | 0.08 | 0.09 | 0.11 |
| Milk type used | 0.07 | 0.08 | 0.10 |
| **C - Lifestyle and environment Total** | **9.70** | **11.54** | **10.72** |
| **D - Physical measurements** |  |  |  |
| Leg fat percentage (right) | 0.68 | 0.81 | 1.79 |
| Waist circumference | 1.24 | 1.47 | 1.44 |
| Impedance of leg (right) | 0.22 | 0.26 | 0.60 |
| Sitting height | 0.43 | 0.51 | 0.54 |
| Body mass index (BMI) | 0.30 | 0.35 | 0.51 |
| Hand grip strength (right) | 0.11 | 0.14 | 0.33 |
| Whole body fat-free mass | 0.11 | 0.13 | 0.25 |
| Impedance of arm (left) | 0.13 | 0.15 | 0.21 |
| Arm fat mass (right) | 0.07 | 0.08 | 0.20 |
| Standing height | 0.13 | 0.16 | 0.18 |
| Hip circumference | 0.10 | 0.12 | 0.18 |
| Weight | 0.06 | 0.07 | 0.16 |
| Contra-indications for spirometry | 0.13 | 0.15 | 0.13 |
| **D - Physical measurements Total** | **3.71** | **4.41** | **6.51** |
| **E - Cognitive function** |  |  |  |
| Mean time to correctly identify matches | 0.57 | 0.68 | 0.96 |
| Time to complete round | 0.55 | 0.65 | 0.63 |
| Number of incorrect matches in round | 0.09 | 0.10 | 0.17 |
| **E - Cognitive function Total** | **1.21** | **1.44** | **1.76** |
| **F - Psychosocial factors** |  |  |  |
| Worry too long after embarrassment | 0.55 | 0.65 | 0.58 |
| Illness, injury, bereavement, stress in last 2 years - Serious illness, injury, or assault of a close relative | 0.20 | 0.24 | 0.30 |
| Risk taking | 0.15 | 0.17 | 0.25 |
| Fed-up feelings | 0.11 | 0.13 | 0.19 |
| Sensitivity / hurt feelings | 0.17 | 0.20 | 0.18 |
| Guilty feelings | 0.08 | 0.09 | 0.17 |
| Irritability | 0.06 | 0.07 | 0.16 |
| Illness, injury, bereavement, stress in last 2 years - serious illness, injury, or assault to yourself | 0.08 | 0.10 | 0.13 |
| Worrier / anxious feelings | 0.09 | 0.11 | 0.09 |
| Illness, injury, bereavement, stress in last 2 years - Death of a close relative | 0.07 | 0.08 | 0.08 |
| **F - Psychosocial factors Total** | **1.56** | **1.85** | **2.14** |
| **G - Self-reported diseases** |  |  |  |
| Number of self-reported cancers | 0.61 | 0.73 | 0.77 |
| Number of self-reported non-cancer illnesses | 0.17 | 0.20 | 0.20 |
| **G - Self-reported diseases Total** | **0.78** | **0.93** | **0.97** |
| **H - Medications & Operations** |  |  |  |
| Number of treatments/medications taken | 0.28 | 0.33 | 0.36 |
| Number of operations, self-reported | 0.05 | 0.06 | 0.06 |
| **H - Medications & Operations Total** | **0.33** | **0.39** | **0.42** |
| **I - Health and medical history** |  |  |  |
| Long-standing illness, disability, or infirmity | 1.51 | 1.80 | 1.44 |
| Overall health rating—poorer | 1.51 | 1.80 | 1.34 |
| Ever had bowel cancer screening | 0.93 | 1.10 | 1.03 |
| Diabetes diagnosed by doctor | 0.49 | 0.59 | 0.50 |
| Vascular/heart problems diagnosed by doctor - heart attack | 0.43 | 0.52 | 0.47 |
| Mother still alive | 0.24 | 0.28 | 0.43 |
| Medication for pain relief, constipation, heartburn - paracetamol | 0.32 | 0.39 | 0.37 |
| Mouth/teeth dental problems - dentures | 0.37 | 0.44 | 0.37 |
| Hearing difficulty/problems with background noise | 0.46 | 0.54 | 0.34 |
| Number of full brothers | 0.27 | 0.33 | 0.29 |
| Pain type(s) experienced in last month - headache | 0.17 | 0.20 | 0.26 |
| Pain type(s) experienced in last month - knee pain | 0.14 | 0.17 | 0.25 |
| Mineral and other dietary supplements - glucosamine | 0.20 | 0.24 | 0.24 |
| Medication for pain relief, constipation, heartburn—none | 0.13 | 0.15 | 0.24 |
| No vascular/heart problems diagnosed by doctor | 0.11 | 0.13 | 0.22 |
| Taking other prescription medications | 0.18 | 0.22 | 0.22 |
| Number of full sisters | 0.10 | 0.12 | 0.21 |
| Current employment status - doing unpaid or voluntary work | 0.17 | 0.20 | 0.20 |
| Father still alive | 0.09 | 0.11 | 0.20 |
| Fractured/broken bones in last 5 years | 0.13 | 0.15 | 0.19 |
| Other serious medical condition/disability diagnosed by doctor | 0.11 | 0.13 | 0.18 |
| Wheeze or whistling in the chest in last year | 0.24 | 0.29 | 0.18 |
| Pain type(s) experienced in last month—none | 0.21 | 0.25 | 0.17 |
| Pain type(s) experienced in last month - neck or shoulder pain | 0.10 | 0.11 | 0.16 |
| Hay fever, allergic rhinitis or eczema diagnosed by doctor | 0.08 | 0.09 | 0.14 |
| Weight change compared with 1 year ago—lost, same, gained | 0.09 | 0.11 | 0.13 |
| Medication for pain relief, constipation, heartburn - ibuprofen (e.g. Nurofen) | 0.06 | 0.07 | 0.12 |
| Hearing difficulty/problems | 0.06 | 0.07 | 0.12 |
| Mouth/teeth dental problems—none | 0.07 | 0.08 | 0.08 |
| Mineral and other dietary supplements—none | 0.10 | 0.11 | 0.07 |
| Mineral and other dietary supplements - fish oil (including cod liver oil) | 0.05 | 0.06 | 0.06 |
| **I - Health and medical history Total** | **9.12** | **10.85** | **10.22** |
| **J - Hospital diagnoses** |  |  |  |
| Sec. - 251.5 palliative care | 8.86 | 10.55 | 9.33 |
| Age at cancer diagnosis | 5.86 | 6.97 | 5.91 |
| Malignant, primary site cancer tumor | 4.29 | 5.10 | 4.23 |
| Sec. - Z51.1 chemotherapy session for neoplasm | 4.29 | 5.10 | 4.20 |
| Sec. - C78.7 secondary malignant neoplasm of liver | 2.71 | 3.23 | 2.87 |
| Sec. - C79.5 Sec. malignant neoplasm of bone and bone marrow | 1.62 | 1.93 | 2.08 |
| Sec. - N17.9 acute renal failure, unspecified | 1.47 | 1.75 | 1.45 |
| Sec. - I10 essential (primary) hypertension | 0.98 | 1.17 | 1.00 |
| Sec. - I48 atrial fibrillation and flutter | 0.87 | 1.04 | 0.85 |
| Histology of cancer tumor - Basal cell carcinoma, NOS | 0.80 | 0.96 | 0.84 |
| Sec. - J90 pleural effusion, not elsewhere classified | 0.84 | 0.99 | 0.80 |
| Reported occurrences of cancer | 0.67 | 0.80 | 0.79 |
| Sec. - Z86.4 personal history of psychoactive substance abuse | 0.77 | 0.92 | 0.74 |
| Sec. - C78.0 secondary malignant neoplasm of lung | 0.46 | 0.55 | 0.65 |
| C61 malignant neoplasm of prostate | 0.41 | 0.48 | 0.56 |
| Sec. - Z51.8 other specified medical care | 0.42 | 0.50 | 0.51 |
| Sec. - I46.9 cardiac arrest, unspecified | 0.42 | 0.50 | 0.45 |
| Sec. - R18 ascites | 0.33 | 0.39 | 0.40 |
| Sec. - J96.9 respiratory failure, unspecified | 0.36 | 0.43 | 0.40 |
| Sec. - C78.6 Sec. malignant neoplasm of retroperitoneum and peritoneum | 0.41 | 0.49 | 0.39 |
| Main - C34.9 bronchus or lung, unspecified | 0.31 | 0.36 | 0.36 |
| Histology of cancer tumor - adenocarcinoma-NOS | 0.17 | 0.20 | 0.35 |
| Behavior of cancer tumor - carcinoma in situ cancer tumor | 0.30 | 0.35 | 0.35 |
| Sec. - N39.0 urinary tract infection, site not specified | 0.35 | 0.41 | 0.33 |
| Sec. - E87.2 acidosis | 0.26 | 0.31 | 0.32 |
| Sec. - J22 unspecified acute lower respiratory infection | 0.26 | 0.31 | 0.30 |
| Main - Z08.0 follow-up exam. after surgery for malignant neoplasm | 0.23 | 0.27 | 0.29 |
| Sec. - F10.2 dependence syndrome | 0.24 | 0.29 | 0.29 |
| Main - C50.9 breast, unspecified | 0.13 | 0.15 | 0.29 |
| Sec. - J18.1 lobar pneumonia, unspecified | 0.26 | 0.31 | 0.28 |
| Main - M17.9 gonarthrosis, unspecified | 0.24 | 0.29 | 0.28 |
| Sec. - J84.1 other interstitial pulmonary diseases with fibrosis | 0.24 | 0.28 | 0.28 |
| Sec. - I95.9 hypotension, unspecified | 0.20 | 0.23 | 0.28 |
| Histology of cancer tumor - duct adenocarcinoma | 0.21 | 0.24 | 0.27 |
| Histology of cancer tumor - glioblastoma-NOS | 0.23 | 0.27 | 0.26 |
| Sec. - R56.8 other and unspecified convulsions | 0.18 | 0.21 | 0.24 |
| Main - G12.2 motor neuron disease | 0.21 | 0.25 | 0.23 |
| Sec. - D64.9 anemia, unspecified | 0.21 | 0.25 | 0.22 |
| Cancer report format | 0.34 | 0.40 | 0.22 |
| Sec. - R63.4 abnormal weight loss | 0.18 | 0.22 | 0.21 |
| Sec. - F10.1 harmful use | 0.18 | 0.21 | 0.20 |
| Sec. - I50.0 congestive heart failure | 0.14 | 0.17 | 0.20 |
| Main - K63.5 polyp of colon | 0.11 | 0.14 | 0.20 |
| Sec. - F32.9 depressive episode, unspecified | 0.07 | 0.09 | 0.19 |
| Main - K21.9 gastro-esophageal reflux disease without esophagitis | 0.12 | 0.14 | 0.19 |
| Sec. - A41.9 septicemia, unspecified | 0.13 | 0.15 | 0.18 |
| Sec. - I50.1 left ventricular failure | 0.16 | 0.19 | 0.18 |
| Main - J18.1 lobar pneumonia, unspecified | 0.17 | 0.21 | 0.18 |
| Sec. - G20 Parkinson's disease | 0.12 | 0.14 | 0.17 |
| Main - R69 unknown and unspecified causes of morbidity | 0.10 | 0.12 | 0.16 |
| Sec. - M17.9 gonarthrosis, unspecified | 0.08 | 0.09 | 0.16 |
| Sec. - I46.0 cardiac arrest with successful resuscitation | 0.15 | 0.18 | 0.16 |
| Main - J90 pleural effusion, not elsewhere classified | 0.09 | 0.11 | 0.15 |
| Main - M16.9 coxarthrosis, unspecified | 0.09 | 0.10 | 0.15 |
| Sec. - F03 unspecified dementia | 0.11 | 0.13 | 0.14 |
| Main - C79.3 Sec. malignant neoplasm of brain and cerebral meninges | 0.09 | 0.10 | 0.13 |
| Main - D64.9 anemia, unspecified | 0.09 | 0.10 | 0.13 |
| Sec. - J69.0 pneumonitis due to food and vomit | 0.09 | 0.11 | 0.13 |
| Histology of cancer tumor - neoplasm | 0.08 | 0.09 | 0.13 |
| Sec. - Z51.3 blood transfusion without reported diagnosis | 0.09 | 0.11 | 0.12 |
| Histology of cancer tumor - carcinoma-NOS | 0.08 | 0.10 | 0.12 |
| Sec. - R41.0 disorientation, unspecified | 0.07 | 0.08 | 0.11 |
| Main - K40.9 unilateral or unspecified inguinal hernia | 0.07 | 0.09 | 0.11 |
| Sec. - R94.5 abnormal results of liver function studies | 0.08 | 0.09 | 0.11 |
| Sec. - E87.1 hypo-osmolality and hyponatremia | 0.07 | 0.09 | 0.11 |
| Main - I61.9 intracerebral hemorrhage, unspecified | 0.11 | 0.13 | 0.10 |
| Main - I63.9 cerebral infarction, unspecified | 0.08 | 0.10 | 0.10 |
| Sec. - Z86.7 personal history of diseases of the circulatory system | 0.11 | 0.13 | 0.10 |
| Sec. - J18.9 pneumonia, unspecified | 0.05 | 0.07 | 0.10 |
| Main - J22 unspecified acute lower respiratory infection | 0.07 | 0.09 | 0.10 |
| Main - C44.3 Other/unspecified skin of other and unspecified parts of face | 0.05 | 0.06 | 0.10 |
| Main - C71.9 brain, unspecified | 0.07 | 0.09 | 0.09 |
| Main - J84.1 other interstitial pulmonary diseases with fibrosis | 0.09 | 0.10 | 0.09 |
| Sec. - G12.2 motor neuron disease | 0.10 | 0.12 | 0.09 |
| Sec. - C79.3 secondary malignant neoplasm of brain and cerebral meninges | 0.06 | 0.08 | 0.08 |
| Main - R91 abnormal findings on diagnostic imaging of lung | 0.06 | 0.08 | 0.08 |
| Sec. - R11 nausea and vomiting | 0.07 | 0.08 | 0.08 |
| Sec. - G30.9 Alzheimer's disease, unspecified | 0.05 | 0.06 | 0.08 |
| Sec. - G91.9 hydrocephalus, unspecified | 0.06 | 0.08 | 0.08 |
| Sec. - E11.9 non-insulin-dependent diabetes mellites complications | 0.06 | 0.07 | 0.08 |
| Sec. - G93.1 anoxic brain damage, not elsewhere classified | 0.05 | 0.06 | 0.07 |
| Main - I60.9 subarachnoid hemorrhage, unspecified | 0.07 | 0.08 | 0.07 |
| Sec. - I63.9 Cerebral infarction, unspecified | 0.06 | 0.07 | 0.07 |
| Main - I21.0 acute transmural myocardial infarction of anterior wall | 0.06 | 0.07 | 0.05 |
| Main - I21.9 acute myocardial infarction, unspecified | 0.06 | 0.07 | 0.05 |
| **J - Hospital diagnoses Total** | **45.62** | **54.28** | **49.61** |
| **Grand Total** | **84.04** | **100.00** | **100.00** |

Abbreviations: A level, advanced level; AS level, advanced subsidiary level; BMI, body mass index; CSE, certificate of secondary education; NOS, not otherwise specified; Sec., secondary diagnosis.

^a^ International classification of diseases (ICD) codes are given for diagnoses. Predictor names are modified by adding additional text after a ‘—’ for some predictors to reflect how higher value(s) are coded.

^b^ SHAP values (normalized for 100%) from a GBDT model with all predictors in its input totaling 84%,

^c^ SHAP values adjusted for a total of 100%

^d^ SHAP values from a GBDT model with only important predictors in its input

**Supplementary Table 4.** Ranking of predictors based on SHAP (SHapley Additive exPlanation) values, hazard ratios (HR) and *P*-values from unadjusted and adjusted Cox models are also reported. The table is sorted on *P*-values from the adjusted Cox models.

| **Category** | **Predictor ID** | **Predictor name** ^a^ | **Ranking based on SHAP values** | **Unadjusted hazard ratio** | **Unadjusted hazard ratio 95%**  **confidence interval lower** | **Unadjusted hazard ratio 95%**  **confidence interval upper** | **Unadjusted Cox *P*-value** | **Adjusted hazard ratio** | **Adjusted hazard ratio 95%**  **confidence interval lower** | **Adjusted hazard ratio 95%**  **confidence interval upper** | **Adjusted Cox *P*-value** |
| --- | --- | --- | --- | --- | --- | --- | --- | --- | --- | --- | --- |
| J - Hospital diagnoses | x41204__Z515 | Sec. - 251.5 palliative care | 1 | 82.74 | 76.08 | 89.98 | 2.17E-2312 | 62.36 | 57.06 | 68.14 | 4.07E-1812 |
| J - Hospital diagnoses | x41204__C787 | Sec. - C78.7 secondary malignant neoplasm of liver | 8 | 65.95 | 59.87 | 72.65 | 2.14E-1566 | 49.91 | 45.19 | 55.13 | 1.60E-1292 |
| J - Hospital diagnoses | x41204__Z511 | Sec. - Z51.1 chemotherapy session for neoplasm | 5 | 18.40 | 17.01 | 19.90 | 1.73E-1155 | 17.02 | 15.70 | 18.44 | 6.28E-1039 |
| J - Hospital diagnoses | x41204__C780 | Sec. - C78.0 secondary malignant neoplasm of lung | 34 | 59.92 | 53.61 | 66.97 | 8.35E-1133 | 47.63 | 42.52 | 53.35 | 3.03E-970 |
| J - Hospital diagnoses | x41204__C795 | Sec. - C79.5 Sec. malignant neoplasm of bone and bone marrow | 9 | 55.88 | 50.05 | 62.38 | 2.65E-1117 | 39.91 | 35.64 | 44.68 | 3.38E-890 |
| J - Hospital diagnoses | x40009 | Reported occurrences of cancer | 26 |  |  |  | 4.04E-806 |  |  |  | 2.66E-677 |
| J - Hospital diagnoses | x40012__3 | Malignant, primary site cancer tumor | 4 | 10.87 | 10.09 | 11.70 | 1.65E-864 | 9.00 | 8.34 | 9.73 | 9.26E-680 |
| J - Hospital diagnoses | x40008 | Age at cancer diagnosis | 2 |  |  |  | 6.69E-863 |  |  |  | 4.00E-662 |
| J - Hospital diagnoses | x40019 | Cancer report format | 91 |  |  |  | 8.37E-770 |  |  |  | 6.14E-646 |
| J - Hospital diagnoses | x41204__C786 | Sec. - C78.6 Sec. malignant neoplasm of retroperitoneum and peritoneum | 50 | 42.02 | 36.48 | 48.41 | 1.19E-584 | 38.61 | 33.40 | 44.62 | 3.28E-534 |
| J - Hospital diagnoses | x41204__J90 | Sec. - J90 pleural effusion, not elsewhere classified | 25 | 18.26 | 16.45 | 20.27 | 3.26E-647 | 12.80 | 11.51 | 14.24 | 8.10E-481 |
| J - Hospital diagnoses | x41202__C349 | Main - C34.9 bronchus or lung, unspecified | 55 | 47.50 | 41.12 | 54.87 | 7.12E-601 | 29.93 | 25.80 | 34.72 | 3.55E-439 |
| J - Hospital diagnoses | x41204__N179 | Sec. - N17.9 acute renal failure, unspecified | 12 | 18.75 | 16.89 | 20.82 | 5.12E-658 | 11.65 | 10.46 | 12.98 | 2.02E-433 |
| J - Hospital diagnoses | x41204__C793 | Sec. - C79.3 secondary malignant neoplasm of brain and cerebral meninges | 177 | 61.13 | 51.52 | 72.55 | 3.26E-484 | 49.74 | 41.78 | 59.23 | 1.86E-420 |
| J - Hospital diagnoses | x41204__R18 | Sec. - R18 ascites | 47 | 25.44 | 21.88 | 29.57 | 7.19E-388 | 19.85 | 17.05 | 23.10 | 2.26E-325 |
| J - Hospital diagnoses | x41204__Z518 | Sec. - Z51.8 other specified medical care | 42 | 33.72 | 28.51 | 39.89 | 1.40E-368 | 26.83 | 22.64 | 31.81 | 2.64E-314 |
| J - Hospital diagnoses | x40011__8140 | Histology of cancer tumor - adenocarcinoma-NOS | 56 | 9.20 | 8.43 | 10.05 | 5.47E-538 | 5.79 | 5.27 | 6.35 | 3.89E-299 |
| J - Hospital diagnoses | x41204__J181 | Sec. - J18.1 lobar pneumonia, unspecified | 71 | 17.80 | 15.55 | 20.37 | 7.69E-382 | 12.10 | 10.55 | 13.88 | 1.65E-279 |
| J - Hospital diagnoses | x41204__A419 | Sec. - A41.9 septicemia, unspecified | 117 | 19.96 | 17.15 | 23.24 | 9.50E-326 | 15.66 | 13.43 | 18.24 | 6.41E-272 |
| J - Hospital diagnoses | x41204__J969 | Sec. - J96.9 respiratory failure, unspecified | 49 | 20.78 | 17.89 | 24.12 | 1.41E-346 | 13.79 | 11.84 | 16.06 | 3.50E-249 |
| A - Baseline characteristics | x21022 | Age at recruitment | 3 | 1.86 | 1.79 | 1.92 | 1.63E-248 | 1.86 | 1.79 | 1.92 | 1.63E-248 |
| J - Hospital diagnoses | x41204__Z513 | Sec. - Z51.3 blood transfusion without reported diagnosis | 155 | 22.33 | 18.91 | 26.38 | 1.58E-292 | 17.34 | 14.65 | 20.52 | 5.54E-242 |
| J - Hospital diagnoses | x41202__J90 | Main - J90 pleural effusion, not elsewhere classified | 138 | 21.90 | 18.69 | 25.66 | 1.73E-318 | 14.99 | 12.76 | 17.60 | 2.75E-238 |
| J - Hospital diagnoses | x41204__D649 | Sec. - D64.9 anemia, unspecified | 88 | 7.87 | 7.07 | 8.76 | 1.00E-300 | 6.26 | 5.61 | 6.99 | 6.59E-236 |
| J - Hospital diagnoses | x41202__C793 | Main - C79.3 Sec. malignant neoplasm of brain and cerebral meninges | 145 | 51.82 | 41.41 | 64.84 | 3.99E-261 | 44.09 | 35.15 | 55.31 | 5.06E-235 |
| J - Hospital diagnoses | x41204__N390 | Sec. - N39.0 urinary tract infection, site not specified | 60 | 7.92 | 7.06 | 8.90 | 3.27E-268 | 6.16 | 5.47 | 6.92 | 3.24E-202 |
| J - Hospital diagnoses | x41204__R11 | Sec. - R11 nausea and vomiting | 181 | 5.91 | 5.28 | 6.62 | 5.14E-210 | 5.56 | 4.96 | 6.23 | 3.31E-191 |
| J - Hospital diagnoses | x41204__E872 | Sec. - E87.2 acidosis | 62 | 22.30 | 18.59 | 26.76 | 7.95E-245 | 15.80 | 13.12 | 19.02 | 1.31E-186 |
| J - Hospital diagnoses | x41204__Z864 | Sec. - Z86.4 personal history of psychoactive substance abuse | 30 | 4.93 | 4.53 | 5.35 | 3.35E-309 | 3.63 | 3.32 | 3.96 | 1.53E-180 |
| J - Hospital diagnoses | x41204__J22 | Sec. - J22 unspecified acute lower respiratory infection | 63 | 9.81 | 8.60 | 11.20 | 8.62E-253 | 6.88 | 6.01 | 7.86 | 1.11E-174 |
| J - Hospital diagnoses | x41204__J189 | Sec. - J18.9 pneumonia, unspecified | 170 | 16.50 | 13.93 | 19.54 | 3.17E-231 | 11.52 | 9.70 | 13.67 | 1.50E-171 |
| J - Hospital diagnoses | x41202__J181 | Main - J18.1 lobar pneumonia, unspecified | 119 | 9.81 | 8.55 | 11.25 | 1.52E-233 | 6.92 | 6.02 | 7.96 | 7.82E-164 |
| G - Self-reported diseases | x134 | Number of self-reported cancers | 29 | 2.79 | 2.62 | 2.97 | 3.45E-222 | 2.49 | 2.33 | 2.66 | 1.04E-163 |
| J - Hospital diagnoses | x41204__Z867 | Sec. - Z86.7 personal history of diseases of the circulatory system | 169 | 5.32 | 4.84 | 5.86 | 6.93E-257 | 3.81 | 3.46 | 4.20 | 1.45E-158 |
| J - Hospital diagnoses | x40011__9440 | Histology of cancer tumor - glioblastoma-NOS | 76 | 56.88 | 43.10 | 75.08 | 3.91E-179 | 45.51 | 34.37 | 60.26 | 1.76E-156 |
| J - Hospital diagnoses | x40011__8000 | Histology of cancer tumor - neoplasm | 154 | 11.55 | 9.73 | 13.70 | 3.88E-173 | 10.93 | 9.16 | 13.06 | 8.27E-154 |
| I - Health and medical history | x2178 | Overall health rating—poorer | 16 |  |  |  | 2.33E-178 |  |  |  | 1.23E-145 |
| J - Hospital diagnoses | x41202__D649 | Main - D64.9 anemia, unspecified | 147 | 7.22 | 6.31 | 8.26 | 2.40E-181 | 5.94 | 5.18 | 6.80 | 3.12E-145 |
| B - Sociodemographics | x6146__100 | Attendance/disability/mobility allowance—none | 31 | 0.23 | 0.21 | 0.25 | 3.31E-232 | 0.30 | 0.28 | 0.33 | 2.02E-143 |
| J - Hospital diagnoses | x41204__I469 | Sec. - I46.9 cardiac arrest, unspecified | 45 | 46.35 | 36.16 | 59.40 | 9.27E-202 | 25.29 | 19.66 | 32.54 | 2.81E-139 |
| F - Psychosocial factors | x6145__1 | Illness, injury, bereavement, stress in last 2 years - serious illness, injury, or assault to yourself | 152 | 3.20 | 2.94 | 3.49 | 1.64E-157 | 2.97 | 2.73 | 3.24 | 1.61E-136 |
| H - Medications & Operations | x137 | Number of treatments/medications taken | 54 | 1.17 | 1.16 | 1.18 | 4.03E-274 | 1.13 | 1.12 | 1.14 | 1.92E-135 |
| J - Hospital diagnoses | x41204__I959 | Sec. - I95.9 hypotension, unspecified | 73 | 9.01 | 7.78 | 10.44 | 9.63E-189 | 6.54 | 5.64 | 7.59 | 8.94E-135 |
| B - Sociodemographics | x6146__2 | Attendance/disability/mobility allowance - disability living allowance | 94 | 4.36 | 3.95 | 4.80 | 4.63E-194 | 3.53 | 3.19 | 3.91 | 2.18E-132 |
| J - Hospital diagnoses | x41204__R410 | Sec. - R41.0 disorientation, unspecified | 161 | 14.47 | 12.21 | 17.16 | 1.59E-207 | 8.65 | 7.27 | 10.29 | 5.16E-131 |
| J - Hospital diagnoses | x41204__I48 | Sec. - I48 atrial fibrillation and flutter | 23 | 6.27 | 5.67 | 6.94 | 1.46E-274 | 3.69 | 3.32 | 4.10 | 1.43E-129 |
| J - Hospital diagnoses | x41204__E871 | Sec. - E87.1 hypo-osmolality and hyponatremia | 165 | 12.65 | 10.58 | 15.12 | 7.79E-171 | 8.71 | 7.27 | 10.43 | 1.35E-122 |
| J - Hospital diagnoses | x41202__J22 | Main - J22 unspecified acute lower respiratory infection | 171 | 7.45 | 6.43 | 8.62 | 2.85E-159 | 5.69 | 4.91 | 6.60 | 3.76E-118 |
| J - Hospital diagnoses | x40011__8010 | Histology of cancer tumor - carcinoma-NOS | 157 | 6.55 | 5.53 | 7.75 | 9.75E-106 | 7.31 | 6.16 | 8.67 | 1.08E-114 |
| J - Hospital diagnoses | x41204__I500 | Sec. - I50.0 congestive heart failure | 101 | 13.26 | 11.23 | 15.65 | 9.77E-205 | 7.12 | 6.01 | 8.43 | 1.45E-113 |
| J - Hospital diagnoses | x41204__R945 | Sec. - R94.5 abnormal results of liver function studies | 162 | 7.62 | 6.43 | 9.02 | 2.41E-122 | 6.66 | 5.62 | 7.90 | 6.92E-106 |
| J - Hospital diagnoses | x41202__C719 | Main - C71.9 brain, unspecified | 174 | 45.97 | 32.44 | 65.15 | 1.14E-102 | 41.62 | 29.24 | 59.25 | 3.79E-95 |
| J - Hospital diagnoses | x41204__I10 | Sec. - I10 essential (primary) hypertension | 21 | 3.24 | 3.01 | 3.48 | 5.90E-217 | 2.22 | 2.06 | 2.40 | 1.63E-91 |
| J - Hospital diagnoses | x41204__I501 | Sec. - I50.1 left ventricular failure | 120 | 9.12 | 7.81 | 10.66 | 2.86E-170 | 5.16 | 4.40 | 6.05 | 3.29E-91 |
| B - Sociodemographics | x6142__4 | Current employment status - unable to work because of sickness or disability | 116 | 3.19 | 2.85 | 3.56 | 3.32E-93 | 3.35 | 2.98 | 3.77 | 2.52E-90 |
| I - Health and medical history | x2188 | Long-standing illness, disability, or infirmity | 14 | 2.71 | 2.51 | 2.92 | 2.45E-152 | 2.17 | 2.01 | 2.34 | 3.22E-90 |
| C - Lifestyle and environment | x924 | Usual walking pace | 19 |  |  |  | 2.17E-145 |  |  |  | 7.29E-88 |
| J - Hospital diagnoses | x41204__J841 | Sec. - J84.1 other interstitial pulmonary diseases with fibrosis | 69 | 16.50 | 13.08 | 20.81 | 1.27E-123 | 10.14 | 8.02 | 12.81 | 1.04E-83 |
| J - Hospital diagnoses | x41202__C509 | Main - C50.9 breast, unspecified | 72 | 3.61 | 3.10 | 4.21 | 2.28E-60 | 4.83 | 4.10 | 5.68 | 1.35E-80 |
| J - Hospital diagnoses | x41204__E119 | Sec. - E11.9 non-insulin-dependent diabetes mellites complications | 183 | 3.94 | 3.57 | 4.35 | 4.75E-162 | 2.62 | 2.37 | 2.90 | 4.65E-77 |
| A - Baseline characteristics | x31 | Sex—male | 6 | 1.95 | 1.81 | 2.10 | 2.76E-71 |  |  |  | 2.76E-71 |
| J - Hospital diagnoses | x41204__J690 | Sec. - J69.0 pneumonitis due to food and vomit | 149 | 21.39 | 16.03 | 28.54 | 2.50E-96 | 13.58 | 10.16 | 18.16 | 2.35E-69 |
| J - Hospital diagnoses | x41202__J841 | Main - J84.1 other interstitial pulmonary diseases with fibrosis | 175 | 22.37 | 16.24 | 30.80 | 1.02E-80 | 17.41 | 12.62 | 24.00 | 4.53E-68 |
| C - Lifestyle and environment | x20116 | Smoking status—never, previous, current | 15 |  |  |  | 6.94E-95 |  |  |  | 2.93E-67 |
| J - Hospital diagnoses | x41204__R568 | Sec. - R56.8 other and unspecified convulsions | 85 | 11.00 | 8.59 | 14.10 | 2.83E-80 | 8.99 | 7.01 | 11.53 | 4.64E-67 |
| J - Hospital diagnoses | x41202__G122 | Main - G12.2 motor neuron disease | 86 | 35.64 | 24.04 | 52.84 | 9.30E-71 | 29.07 | 19.57 | 43.19 | 1.48E-62 |
| J - Hospital diagnoses | x41204__R634 | Sec. - R63.4 abnormal weight loss | 99 | 5.11 | 4.33 | 6.01 | 8.81E-85 | 4.05 | 3.44 | 4.78 | 3.65E-62 |
| J - Hospital diagnoses | x41204__F102 | Sec. - F10.2 dependence syndrome | 67 | 7.98 | 6.46 | 9.85 | 6.80E-83 | 6.16 | 4.97 | 7.64 | 1.11E-61 |
| I - Health and medical history | x2492 | Taking other prescription medications | 92 | 2.43 | 2.25 | 2.62 | 1.40E-118 | 1.92 | 1.77 | 2.08 | 5.29E-61 |
| J - Hospital diagnoses | x41204__I460 | Sec. - I46.0 cardiac arrest with successful resuscitation | 133 | 18.20 | 13.64 | 24.28 | 1.30E-86 | 11.29 | 8.45 | 15.09 | 2.68E-60 |
| C - Lifestyle and environment | x1239 | Current tobacco smoking—more frequently | 10 |  |  |  | 1.56E-60 |  |  |  | 2.37E-59 |
| J - Hospital diagnoses | x41202__R91 | Main - R91 abnormal findings on diagnostic imaging of lung | 179 | 9.38 | 7.53 | 11.68 | 7.80E-89 | 6.16 | 4.94 | 7.68 | 1.54E-58 |
| G - Self-reported diseases | x135 | Number of self-reported non-cancer illnesses | 102 | 1.19 | 1.17 | 1.21 | 2.81E-139 | 1.14 | 1.12 | 1.15 | 3.92E-58 |
| J - Hospital diagnoses | x41204__G122 | Sec. - G12.2 motor neuron disease | 176 | 31.07 | 20.96 | 46.07 | 1.40E-65 | 23.66 | 15.91 | 35.20 | 6.02E-55 |
| J - Hospital diagnoses | x41202__R69 | Main - R69 unknown and unspecified causes of morbidity | 132 | 3.30 | 2.87 | 3.80 | 3.26E-62 | 3.05 | 2.65 | 3.52 | 3.31E-53 |
| J - Hospital diagnoses | x40011__8500 | Histology of cancer tumor - duct adenocarcinoma | 74 | 2.54 | 2.18 | 2.96 | 3.06E-33 | 3.37 | 2.87 | 3.96 | 6.89E-50 |
| I - Health and medical history | x2473 | Other serious medical condition/disability diagnosed by doctor | 118 | 2.12 | 1.96 | 2.29 | 2.76E-74 | 1.83 | 1.69 | 1.97 | 4.82E-49 |
| J - Hospital diagnoses | x41204__F329 | Sec. - F32.9 depressive episode, unspecified | 109 | 2.87 | 2.50 | 3.30 | 3.80E-50 | 2.85 | 2.48 | 3.28 | 1.43E-48 |
| J - Hospital diagnoses | x41204__F101 | Sec. - F10.1 harmful use | 103 | 5.93 | 4.89 | 7.20 | 4.56E-73 | 4.30 | 3.53 | 5.23 | 5.22E-48 |
| B - Sociodemographics | x6142__1 | In paid employment or self-employed | 7 | 0.32 | 0.30 | 0.35 | 7.72E-174 | 0.52 | 0.47 | 0.57 | 3.83E-44 |
| D - Physical measurements | x47 | Hand grip strength (right) | 61 | 0.92 | 0.89 | 0.96 | 1.00E-05 | 0.71 | 0.67 | 0.74 | 2.29E-43 |
| I - Health and medical history | x2316 | Wheeze or whistling in the chest in last year | 121 | 1.98 | 1.83 | 2.14 | 4.27E-62 | 1.76 | 1.62 | 1.90 | 1.26E-42 |
| J - Hospital diagnoses | x41204__G309 | Sec. - G30.9 Alzheimer's disease, unspecified | 180 | 15.97 | 11.39 | 22.39 | 4.86E-58 | 7.73 | 5.49 | 10.87 | 6.83E-32 |
| I - Health and medical history | x6152__6 | Current employment status - doing unpaid or voluntary work | 107 | 3.88 | 3.33 | 4.53 | 9.12E-67 | 2.54 | 2.17 | 2.97 | 1.16E-31 |
| I - Health and medical history | x6150__1 | Vascular/heart problems diagnosed by doctor - heart attack | 44 | 3.91 | 3.43 | 4.45 | 1.30E-92 | 2.18 | 1.91 | 2.50 | 3.55E-30 |
| A - Baseline characteristics | x189 | Townsend deprivation index | 77 | 1.23 | 1.19 | 1.28 | 4.96E-30 | 1.23 | 1.19 | 1.28 | 4.96E-30 |
| I - Health and medical history | x2443 | Diabetes diagnosed by doctor | 43 | 2.84 | 2.55 | 3.16 | 1.08E-63 | 1.97 | 1.76 | 2.19 | 2.04E-29 |
| C - Lifestyle and environment | x6164__2 | Physical activity in last 4 weeks - other exercises (egg: swimming, cycling, keep fit, bowling) | 32 | 0.55 | 0.51 | 0.59 | 1.74E-52 | 0.65 | 0.60 | 0.70 | 4.02E-27 |
| J - Hospital diagnoses | x41202__I639 | Main - I63.9 cerebral infarction, unspecified | 168 | 5.44 | 4.34 | 6.81 | 2.98E-49 | 3.45 | 2.75 | 4.33 | 6.87E-27 |
| J - Hospital diagnoses | x41204__G931 | Sec. - G93.1 anoxic brain damage, not elsewhere classified | 186 | 32.03 | 18.95 | 54.16 | 2.72E-38 | 18.04 | 10.59 | 30.70 | 1.67E-26 |
| D - Physical measurements | x3088 | Contra-indications for spirometry | 153 |  |  |  | 1.10E-45 |  |  |  | 2.80E-26 |
| H - Medications & Operations | x136 | Number of operations, self-reported | 190 | 1.14 | 1.12 | 1.16 | 1.79E-35 | 1.13 | 1.10 | 1.15 | 9.16E-26 |
| B - Sociodemographics | x728 | No. of vehicles in household | 18 | 0.67 | 0.64 | 0.70 | 5.08E-67 | 0.76 | 0.72 | 0.80 | 1.01E-25 |
| J - Hospital diagnoses | x41204__F03 | Sec. - F03 unspecified dementia | 144 | 12.27 | 8.71 | 17.30 | 1.59E-46 | 6.16 | 4.36 | 8.71 | 7.57E-25 |
| D - Physical measurements | x48 | Waist circumference | 13 | 1.45 | 1.40 | 1.50 | 2.26E-89 | 1.23 | 1.18 | 1.28 | 8.72E-22 |
| I - Health and medical history | x6150__100 | No vascular/heart problems diagnosed by doctor | 90 | 0.48 | 0.45 | 0.52 | 1.21E-86 | 0.70 | 0.65 | 0.75 | 9.17E-21 |
| J - Hospital diagnoses | x41204__I639 | Sec. - I63.9 Cerebral infarction, unspecified | 188 | 12.11 | 8.10 | 18.09 | 4.67E-34 | 6.70 | 4.47 | 10.03 | 2.62E-20 |
| J - Hospital diagnoses | x41204__G919 | Sec. - G91.9 hydrocephalus, unspecified | 182 | 9.71 | 6.11 | 15.43 | 7.07E-22 | 8.76 | 5.51 | 13.95 | 5.58E-20 |
| C - Lifestyle and environment | x20160 | Ever smoked | 66 | 1.67 | 1.55 | 1.81 | 1.49E-38 | 1.43 | 1.32 | 1.55 | 1.86E-18 |
| I - Health and medical history | x6149__6 | Mouth/teeth dental problems - dentures | 53 | 2.26 | 2.09 | 2.44 | 9.05E-92 | 1.44 | 1.33 | 1.57 | 8.54E-18 |
| J - Hospital diagnoses | x41202__I619 | Main - I61.9 intracerebral hemorrhage, unspecified | 166 | 9.89 | 6.50 | 15.04 | 9.47E-27 | 6.21 | 4.08 | 9.46 | 1.69E-17 |
| C - Lifestyle and environment | x943 | Frequency of stair climbing in last 4 weeks | 40 |  |  |  | 4.92E-38 |  |  |  | 2.83E-17 |
| E - Cognitive function | x20023 | Mean time to correctly identify matches | 22 | 1.37 | 1.32 | 1.42 | 9.17E-64 | 1.18 | 1.13 | 1.23 | 1.28E-16 |
| B - Sociodemographics | x709 | Number in household | 17 | 0.70 | 0.67 | 0.73 | 3.82E-76 | 0.85 | 0.81 | 0.88 | 7.43E-16 |
| C - Lifestyle and environment | x2267 | Use of sun/UV protection—always | 33 |  |  |  | 3.59E-41 |  |  |  | 6.22E-15 |
| I - Health and medical history | x1835 | Mother still alive | 46 | 0.38 | 0.35 | 0.42 | 3.86E-114 | 0.68 | 0.62 | 0.75 | 2.05E-14 |
| C - Lifestyle and environment | x1448 | Bread type | 98 |  |  |  | 5.86E-27 |  |  |  | 6.65E-14 |
| J - Hospital diagnoses | x41204__G20 | Sec. - G20 Parkinson's disease | 125 | 5.42 | 3.95 | 7.43 | 1.07E-25 | 3.32 | 2.42 | 4.56 | 1.17E-13 |
| I - Health and medical history | x6154__100 | Medication for pain relief, constipation, heartburn—none | 83 | 0.67 | 0.62 | 0.72 | 3.28E-27 | 0.75 | 0.70 | 0.81 | 1.34E-13 |
| F - Psychosocial factors | x1960 | Fed-up feelings | 115 | 1.16 | 1.08 | 1.25 | 1.04E-04 | 1.32 | 1.22 | 1.42 | 9.28E-13 |
| C - Lifestyle and environment | x6164__5 | Types of physical activity in last 4 weeks - Heavy DIY (egg: weeding, lawn mowing, carpentry, digging) | 105 | 0.83 | 0.77 | 0.90 | 2.56E-06 | 0.75 | 0.69 | 0.81 | 1.53E-12 |
| D - Physical measurements | x20015 | Sitting height | 39 | 0.98 | 0.95 | 1.02 | 2.97E-01 | 0.85 | 0.81 | 0.89 | 6.14E-11 |
| J - Hospital diagnoses | x40006__C61 | C61 malignant neoplasm of prostate | 38 | 3.46 | 2.95 | 4.05 | 1.50E-52 | 1.71 | 1.45 | 2.01 | 1.73E-10 |
| J - Hospital diagnoses | x41202__I609 | Main - I60.9 subarachnoid hemorrhage, unspecified | 187 | 5.45 | 3.28 | 9.05 | 5.73E-11 | 4.91 | 2.95 | 8.16 | 8.32E-10 |
| I - Health and medical history | x1797 | Father still alive | 108 | 0.33 | 0.29 | 0.38 | 9.53E-86 | 0.67 | 0.58 | 0.76 | 1.59E-09 |
| C - Lifestyle and environment | x1478 | Salt added to food | 141 |  |  |  | 5.87E-15 |  |  |  | 6.51E-09 |
| I - Health and medical history | x2306 | Weight change compared with 1 year ago—lost, same, gained | 151 |  |  |  | 2.05E-05 |  |  |  | 2.80E-08 |
| C - Lifestyle and environment | x1349 | Processed meat intake | 59 |  |  |  | 5.13E-23 |  |  |  | 5.19E-08 |
| C - Lifestyle and environment | x1418 | Milk type used | 167 |  |  |  | 2.63E-13 |  |  |  | 6.65E-08 |
| B - Sociodemographics | x670 | Type of accommodation lived in—house, flat, mobile, sheltered, care home | 148 |  |  |  | 1.13E-20 |  |  |  | 9.90E-08 |
| E - Cognitive function | x400 | Time to complete round | 35 | 1.33 | 1.28 | 1.38 | 7.58E-51 | 1.11 | 1.07 | 1.16 | 1.01E-07 |
| I - Health and medical history | x6149__100 | Mouth/teeth dental problems—none | 184 | 0.69 | 0.64 | 0.75 | 6.05E-23 | 0.82 | 0.76 | 0.88 | 1.13E-07 |
| I - Health and medical history | x6154__3 | Medication for pain relief, constipation, heartburn - paracetamol | 52 | 1.14 | 1.05 | 1.24 | 2.25E-03 | 1.26 | 1.16 | 1.37 | 1.43E-07 |
| C - Lifestyle and environment | x6164__4 | Light DIY physical activity in last 4 weeks | 84 | 0.76 | 0.71 | 0.82 | 2.28E-13 | 0.82 | 0.76 | 0.88 | 1.61E-07 |
| C - Lifestyle and environment | x1488 | Tea intake | 51 |  |  |  | 1.07E-07 |  |  |  | 2.41E-07 |
| C - Lifestyle and environment | x6144__5 | Eat eggs, diary, wheat, sugar | 95 | 0.67 | 0.62 | 0.73 | 3.03E-23 | 0.81 | 0.75 | 0.88 | 3.46E-07 |
| J - Hospital diagnoses | x41202__I219 | Main - I21.9 acute myocardial infarction, unspecified | 192 | 2.90 | 2.28 | 3.68 | 2.64E-18 | 1.85 | 1.45 | 2.35 | 5.48E-07 |
| C - Lifestyle and environment | x6164__3 | Types of physical activity in last 4 weeks - strenuous sports | 140 | 0.50 | 0.43 | 0.59 | 2.44E-16 | 0.65 | 0.55 | 0.77 | 5.62E-07 |
| I - Health and medical history | x6179__2 | Mineral and other dietary supplements - glucosamine | 82 | 0.84 | 0.76 | 0.92 | 4.12E-04 | 0.78 | 0.71 | 0.87 | 1.46E-06 |
| B - Sociodemographics | x6138__2 | Qualifications - A levels/AS levels or equivalent | 146 | 0.59 | 0.54 | 0.65 | 3.48E-26 | 0.79 | 0.71 | 0.87 | 1.54E-06 |
| C - Lifestyle and environment | x864 | Number of days/week walked 10+ minutes | 126 | 0.98 | 0.97 | 1.00 | 9.14E-02 | 0.96 | 0.94 | 0.97 | 2.93E-06 |
| D - Physical measurements | x23120 | Arm fat mass (right) | 106 | 1.04 | 1.00 | 1.08 | 3.83E-02 |  |  |  | 3.24E-06 |
| J - Hospital diagnoses | x41202__I210 | Main - I21.0 acute transmural myocardial infarction of anterior wall | 191 | 3.53 | 2.53 | 4.93 | 1.17E-13 | 2.19 | 1.57 | 3.07 | 4.15E-06 |
| J - Hospital diagnoses | x40012__2 | Behavior of cancer tumor - carcinoma in situ cancer tumor | 57 | 1.41 | 1.17 | 1.72 | 4.35E-04 | 1.56 | 1.28 | 1.89 | 7.49E-06 |
| J - Hospital diagnoses | x41202__Z080 | Main - Z08.0 follow-up exam. after surgery for malignant neoplasm | 65 | 2.70 | 2.14 | 3.41 | 8.77E-17 | 1.70 | 1.34 | 2.15 | 9.83E-06 |
| C - Lifestyle and environment | x1289 | Cooked vegetable intake | 89 |  |  |  | 8.83E-08 |  |  |  | 1.38E-05 |
| I - Health and medical history | x6179__1 | Mineral and other dietary supplements - fish oil (including cod liver oil) | 189 | 0.98 | 0.91 | 1.06 | 6.89E-01 | 0.84 | 0.78 | 0.91 | 1.68E-05 |
| B - Sociodemographics | x21000 | Ethnic background | 28 |  |  |  | 1.87E-08 |  |  |  | 1.69E-05 |
| I - Health and medical history | x6159__100 | Pain type(s) experienced in last month—none | 130 | 0.85 | 0.79 | 0.91 | 1.99E-05 | 0.86 | 0.79 | 0.92 | 7.29E-05 |
| D - Physical measurements | x21001 | Body mass index (BMI) | 41 | 1.16 | 1.12 | 1.21 | 4.64E-16 | 1.08 | 1.03 | 1.12 | 2.30E-04 |
| I - Health and medical history | x6152__9 | Hay fever, allergic rhinitis or eczema diagnosed by doctor | 142 | 0.68 | 0.62 | 0.75 | 4.38E-15 | 0.83 | 0.76 | 0.92 | 2.35E-04 |
| B - Sociodemographics | x6142__5 | Current employment status - unemployed | 193 | 1.31 | 1.03 | 1.66 | 2.54E-02 | 1.54 | 1.21 | 1.96 | 4.41E-04 |
| C - Lifestyle and environment | x1369 | Beef intake | 78 |  |  |  | 6.33E-10 |  |  |  | 6.83E-04 |
| I - Health and medical history | x2463 | Fractured/broken bones in last 5 years | 112 | 1.18 | 1.05 | 1.32 | 6.64E-03 | 1.23 | 1.09 | 1.38 | 7.29E-04 |
| C - Lifestyle and environment | x1717 | Skin color—less fair | 135 |  |  |  | 3.70E-06 |  |  |  | 7.79E-04 |
| B - Sociodemographics | x6139__3 | Gas or solid-fuel cooking/heating - an open solid fuel fire that you use regularly in winter time | 114 | 0.68 | 0.58 | 0.79 | 7.87E-07 | 0.78 | 0.67 | 0.91 | 1.74E-03 |
| C - Lifestyle and environment | x1389 | Pork intake | 113 |  |  |  | 3.39E-08 |  |  |  | 1.78E-03 |
| I - Health and medical history | x6179__100 | Mineral and other dietary supplements—none | 185 | 1.00 | 0.93 | 1.07 | 9.30E-01 | 1.13 | 1.05 | 1.21 | 1.82E-03 |
| D - Physical measurements | x23111 | Leg fat percentage (right) | 11 | 0.87 | 0.84 | 0.90 | 1.64E-13 | 1.10 | 1.04 | 1.17 | 1.95E-03 |
| B - Sociodemographics | x6138__4 | Qualifications - CSEs or equivalent | 136 | 0.45 | 0.39 | 0.52 | 7.57E-25 | 0.78 | 0.67 | 0.92 | 2.49E-03 |
| I - Health and medical history | x1883 | Number of full sisters | 96 | 1.06 | 1.03 | 1.09 | 2.28E-05 | 1.04 | 1.02 | 1.07 | 2.54E-03 |
| D - Physical measurements | x50 | Standing height | 122 | 1.10 | 1.06 | 1.14 | 5.64E-07 | 0.93 | 0.88 | 0.98 | 4.35E-03 |
| J - Hospital diagnoses | x41202__M179 | Main - M17.9 gonarthrosis, unspecified | 70 | 0.99 | 0.78 | 1.25 | 9.33E-01 | 0.71 | 0.56 | 0.90 | 5.20E-03 |
| C - Lifestyle and environment | x1518 | Hot drink temperature—less hot | 164 |  |  |  | 9.41E-04 |  |  |  | 8.35E-03 |
| C - Lifestyle and environment | x1359 | Poultry intake | 48 |  |  |  | 3.06E-12 |  |  |  | 9.20E-03 |
| I - Health and medical history | x2257 | Hearing difficulty/problems with background noise | 58 | 1.39 | 1.29 | 1.50 | 2.94E-18 | 1.10 | 1.02 | 1.19 | 9.56E-03 |
| D - Physical measurements | x49 | Hip circumference | 124 | 1.08 | 1.04 | 1.12 | 6.53E-05 | 1.05 | 1.01 | 1.09 | 1.02E-02 |
| D - Physical measurements | x21002 | Weight | 137 | 1.20 | 1.16 | 1.24 | 9.56E-23 | 1.06 | 1.01 | 1.10 | 1.04E-02 |
| D - Physical measurements | x23110 | Impedance of arm (left) | 97 | 0.81 | 0.78 | 0.84 | 6.08E-28 | 1.07 | 1.02 | 1.13 | 1.15E-02 |
| J - Hospital diagnoses | x41202__M169 | Main - M16.9 coxarthrosis, unspecified | 139 | 0.99 | 0.74 | 1.31 | 9.25E-01 | 0.71 | 0.53 | 0.94 | 1.61E-02 |
| F - Psychosocial factors | x2000 | Worry too long after embarrassment | 37 | 0.79 | 0.73 | 0.85 | 8.60E-10 | 0.91 | 0.85 | 0.98 | 1.76E-02 |
| C - Lifestyle and environment | x1647 | Country of birth (UK/elsewhere) | 150 |  |  |  | 1.74E-06 |  |  |  | 1.87E-02 |
| B - Sociodemographics | x6139__100 | Gas or solid-fuel cooking/heating—none | 159 | 1.28 | 1.17 | 1.40 | 7.30E-08 | 1.11 | 1.02 | 1.22 | 2.15E-02 |
| D - Physical measurements | x23107 | Impedance of leg (right) | 36 | 0.81 | 0.78 | 0.84 | 7.55E-28 | 0.95 | 0.92 | 0.99 | 2.32E-02 |
| I - Health and medical history | x2345 | Ever had bowel cancer screening | 20 | 1.64 | 1.52 | 1.76 | 8.11E-37 | 1.10 | 1.01 | 1.19 | 2.39E-02 |
| I - Health and medical history | x6159__3 | Pain type(s) experienced in last month - neck or shoulder pain | 131 | 1.11 | 1.02 | 1.20 | 1.81E-02 | 1.10 | 1.01 | 1.19 | 2.88E-02 |
| C - Lifestyle and environment | x1697 | Comparative height size at age 10 | 100 |  |  |  | 1.58E-01 |  |  |  | 6.66E-02 |
| F - Psychosocial factors | x2040 | Risk taking | 79 | 1.11 | 1.03 | 1.21 | 1.03E-02 | 1.08 | 0.99 | 1.18 | 6.84E-02 |
| F - Psychosocial factors | x6145__2 | Illness, injury, bereavement, stress in last 2 years - Serious illness, injury, or assault of a close relative | 64 | 0.72 | 0.63 | 0.82 | 1.23E-06 | 0.89 | 0.78 | 1.01 | 7.48E-02 |
| J - Hospital diagnoses | x41202__K409 | Main - K40.9 unilateral or unspecified inguinal hernia | 163 | 1.43 | 1.20 | 1.70 | 6.70E-05 | 0.85 | 0.71 | 1.02 | 8.12E-02 |
| B - Sociodemographics | x699 | Length of time at current address | 27 | 1.26 | 1.21 | 1.30 | 5.80E-34 | 1.03 | 0.99 | 1.07 | 9.90E-02 |
| J - Hospital diagnoses | x41204__M179 | Sec. - M17.9 gonarthrosis, unspecified | 128 | 1.43 | 1.14 | 1.79 | 1.83E-03 | 1.21 | 0.96 | 1.52 | 9.97E-02 |
| F - Psychosocial factors | x6145__3 | Illness, injury, bereavement, stress in last 2 years - Death of a close relative | 178 | 0.88 | 0.80 | 0.97 | 7.48E-03 | 0.93 | 0.85 | 1.02 | 1.22E-01 |
| I - Health and medical history | x2247 | Hearing difficulty/problems | 160 |  |  |  | 5.00E-18 |  |  |  | 1.36E-01 |
| A - Baseline characteristics | x52 | Month of birth | 87 |  |  |  | 1.81E-01 |  |  |  | 1.81E-01 |
| I - Health and medical history | x6154__2 | Medication for pain relief, constipation, heartburn - ibuprofen (e.g. Nurofen) | 158 | 0.71 | 0.63 | 0.80 | 1.26E-08 | 0.93 | 0.82 | 1.04 | 2.05E-01 |
| I - Health and medical history | x6159__1 | Pain type(s) experienced in last month - headache | 75 | 0.72 | 0.65 | 0.79 | 4.35E-11 | 0.94 | 0.85 | 1.04 | 2.08E-01 |
| J - Hospital diagnoses | x41202__K219 | Main - K21.9 gastro-esophageal reflux disease without esophagitis | 111 | 1.27 | 0.96 | 1.67 | 9.17E-02 | 1.19 | 0.91 | 1.58 | 2.10E-01 |
| B - Sociodemographics | x6139__2 | Gas or solid-fuel cooking/heating - a gas fire that you use regularly in winter time | 110 | 0.99 | 0.92 | 1.07 | 7.81E-01 | 0.95 | 0.88 | 1.03 | 2.14E-01 |
| I - Health and medical history | x1873 | Number of full brothers | 68 | 1.04 | 1.01 | 1.07 | 4.14E-03 | 1.02 | 0.99 | 1.04 | 2.25E-01 |
| I - Health and medical history | x6159__7 | Pain type(s) experienced in last month - knee pain | 81 | 1.19 | 1.10 | 1.30 | 4.24E-05 | 1.05 | 0.97 | 1.15 | 2.33E-01 |
| F - Psychosocial factors | x1940 | Irritability | 134 | 1.00 | 0.92 | 1.09 | 9.58E-01 | 1.05 | 0.97 | 1.14 | 2.55E-01 |
| J - Hospital diagnoses | x41202__C443 | Main - C44.3 Other/unspecified malignant neoplasm of skin of other and unspecified parts of face | 172 | 1.21 | 0.90 | 1.63 | 2.10E-01 | 0.84 | 0.63 | 1.14 | 2.61E-01 |
| F - Psychosocial factors | x2030 | Guilty feelings | 129 | 0.84 | 0.77 | 0.91 | 4.06E-05 | 0.95 | 0.88 | 1.04 | 2.71E-01 |
| F - Psychosocial factors | x1950 | Sensitivity / hurt feelings | 123 | 0.90 | 0.84 | 0.97 | 7.37E-03 | 1.04 | 0.97 | 1.12 | 2.91E-01 |
| J - Hospital diagnoses | x41202__K635 | Main - K63.5 polyp of colon | 104 | 1.50 | 1.21 | 1.86 | 1.92E-04 | 1.10 | 0.88 | 1.36 | 4.03E-01 |
| C - Lifestyle and environment | x1408 | Cheese intake | 143 |  |  |  | 5.68E-02 |  |  |  | 5.38E-01 |
| C - Lifestyle and environment | x1438 | Bread intake | 93 | 1.13 | 1.09 | 1.17 | 1.27E-10 | 0.99 | 0.95 | 1.03 | 6.64E-01 |
| D - Physical measurements | x23101 | Whole body fat-free mass | 80 | 1.20 | 1.16 | 1.24 | 5.55E-22 | 1.01 | 0.95 | 1.07 | 7.26E-01 |
| C - Lifestyle and environment | x1707 | Handedness—right-handed, left-handed, use both hands | 156 |  |  |  | 5.12E-01 |  |  |  | 7.97E-01 |
| F - Psychosocial factors | x1980 | Worrier / anxious feelings | 173 | 0.88 | 0.82 | 0.95 | 1.22E-03 | 1.01 | 0.93 | 1.08 | 8.92E-01 |
| E - Cognitive function | x399 | Number of incorrect matches in round | 127 | 1.10 | 1.06 | 1.14 | 2.27E-07 | 1.00 | 0.97 | 1.04 | 9.08E-01 |
| J - Hospital diagnoses | x40011__8090 | Histology of cancer tumor - Basal cell carcinoma, NOS | 24 | 1.41 | 1.18 | 1.68 | 1.38E-04 | 1.00 | 0.84 | 1.20 | 9.58E-01 |

Abbreviations: A level, advanced level; AS level, advanced subsidiary level; CSE, certificate of secondary education; NOS, not otherwise specified; Sec., secondary diagnosis.

^a^ International classification of diseases (ICD) codes are given for diagnoses. Predictor names are modified by adding additional text after a ‘—’ for some predictors to reflect how higher value(s) are coded.

^b^ adjusted for age, sex, Townsend deprivation index, assessment center and month of birth.

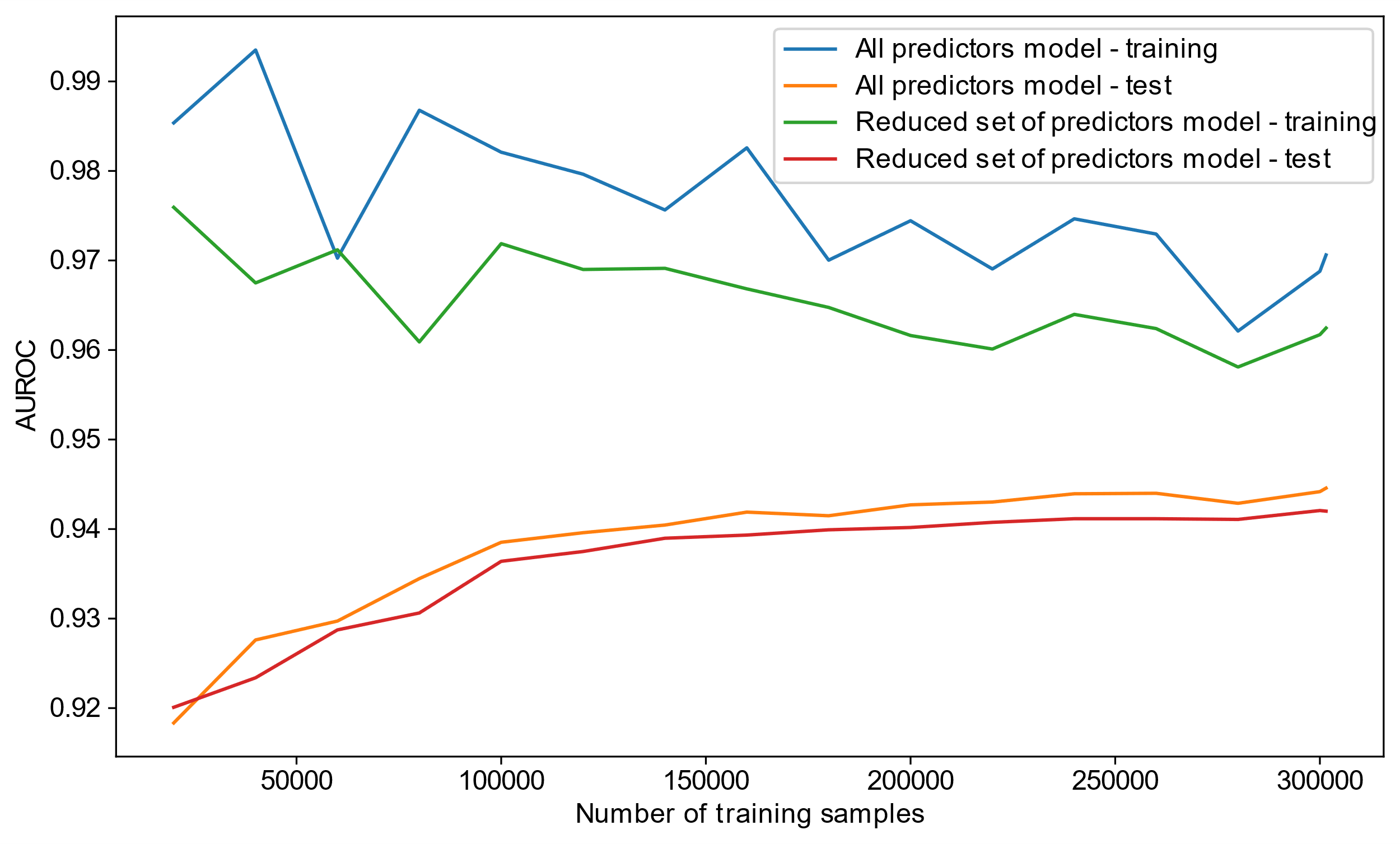

**Supplementary Figure 1**. Learning curve analysis showing performance of set of all predictors model and reduced set of predictors models on training set and test set with increasing number of samples used for training. Model performance was assessed using area under the receiver operating characteristics curve (AUROC).

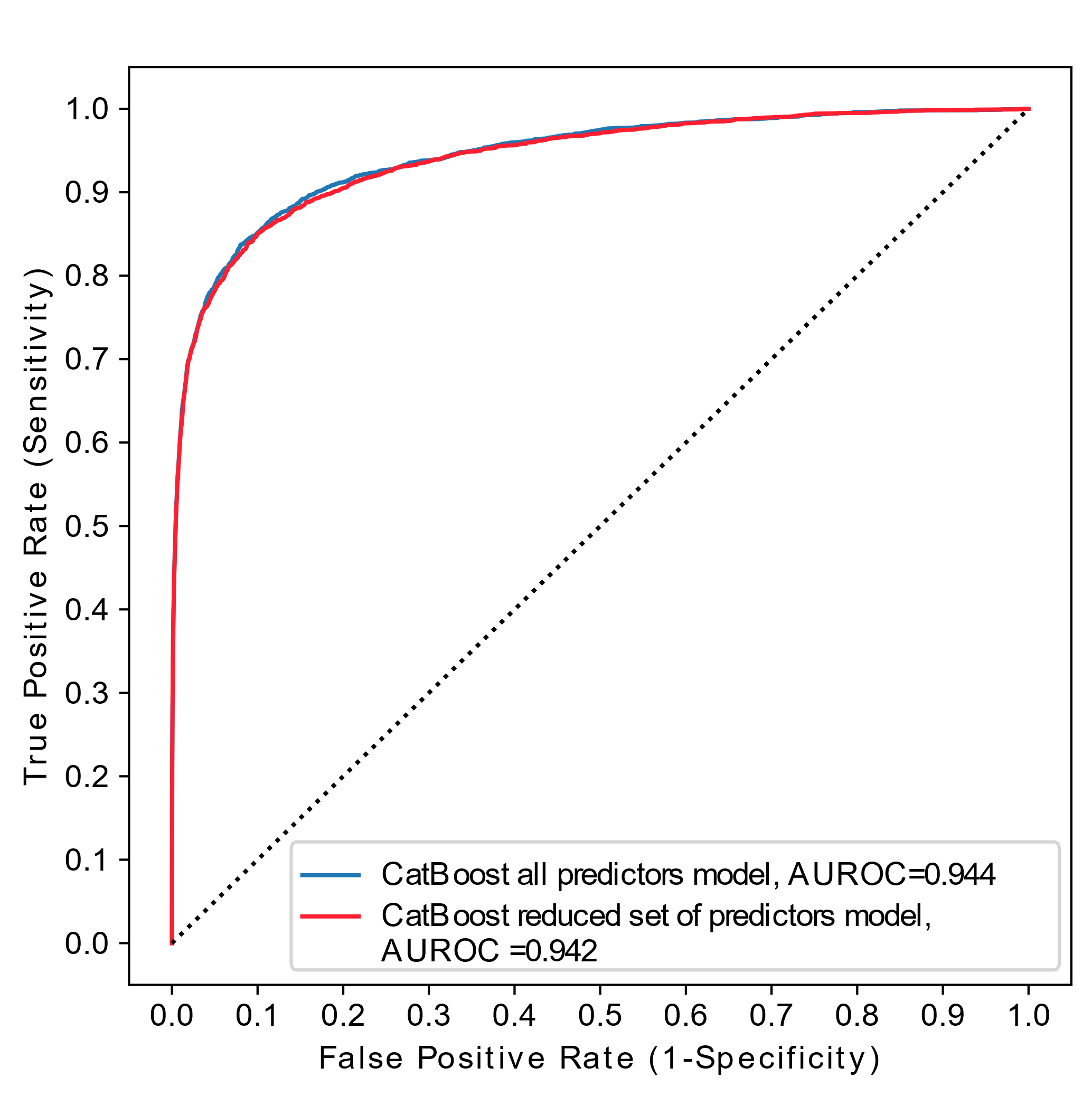

**Supplementary Figure 2**. Receiver operating characteristics (ROC) curves for gradient boosting decision trees (GBDT) models with all predictors and with the 193 important predictors.

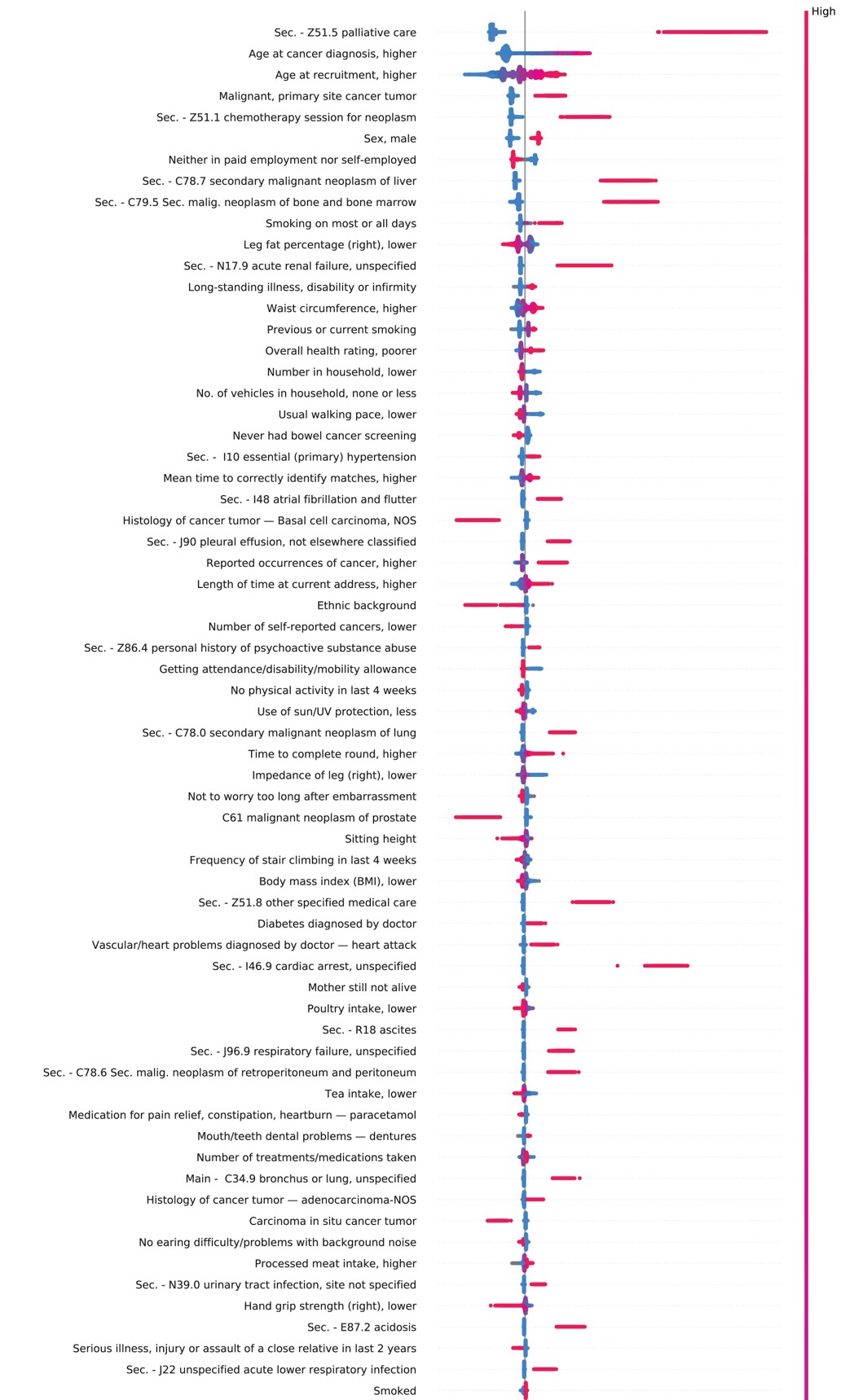

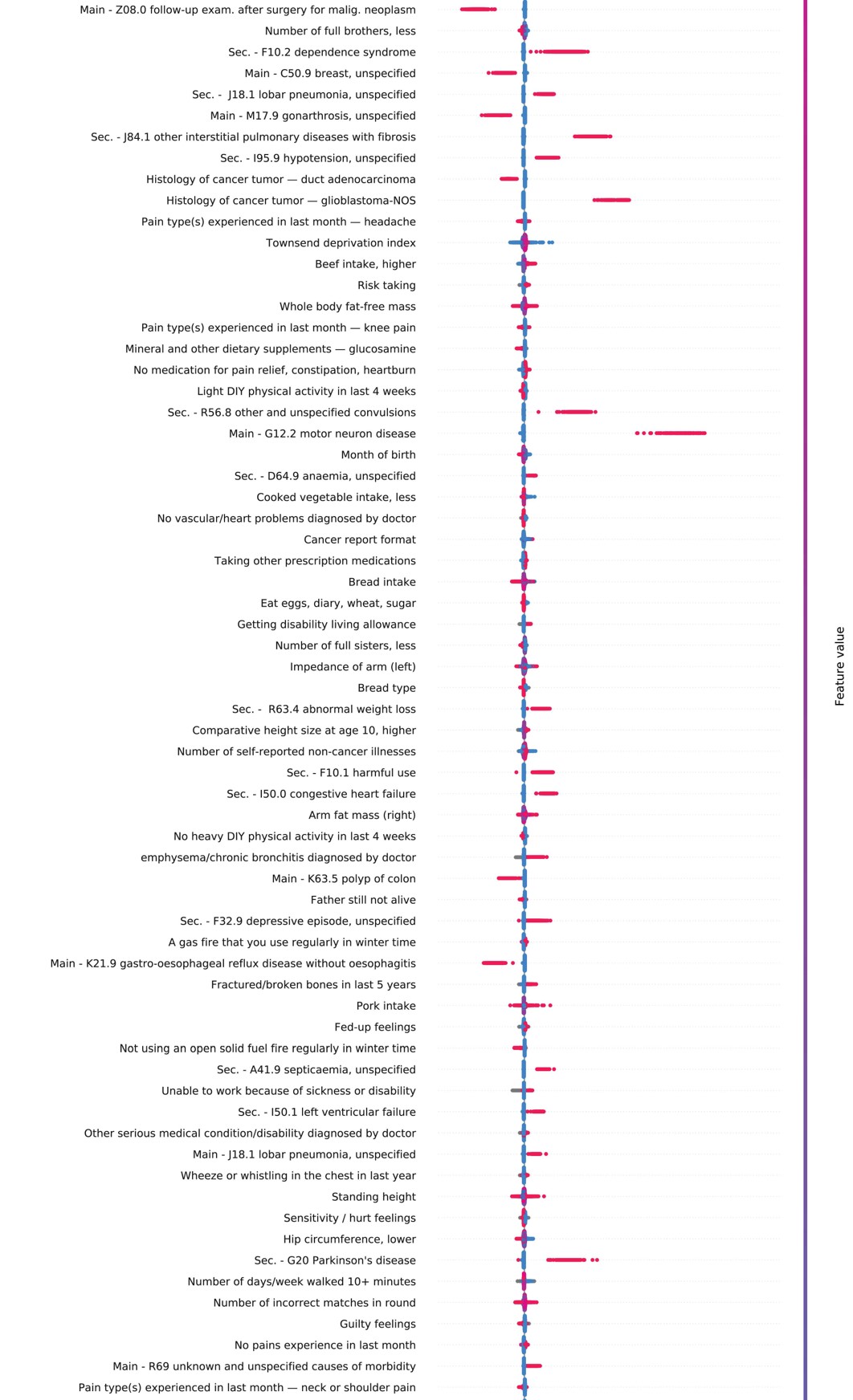

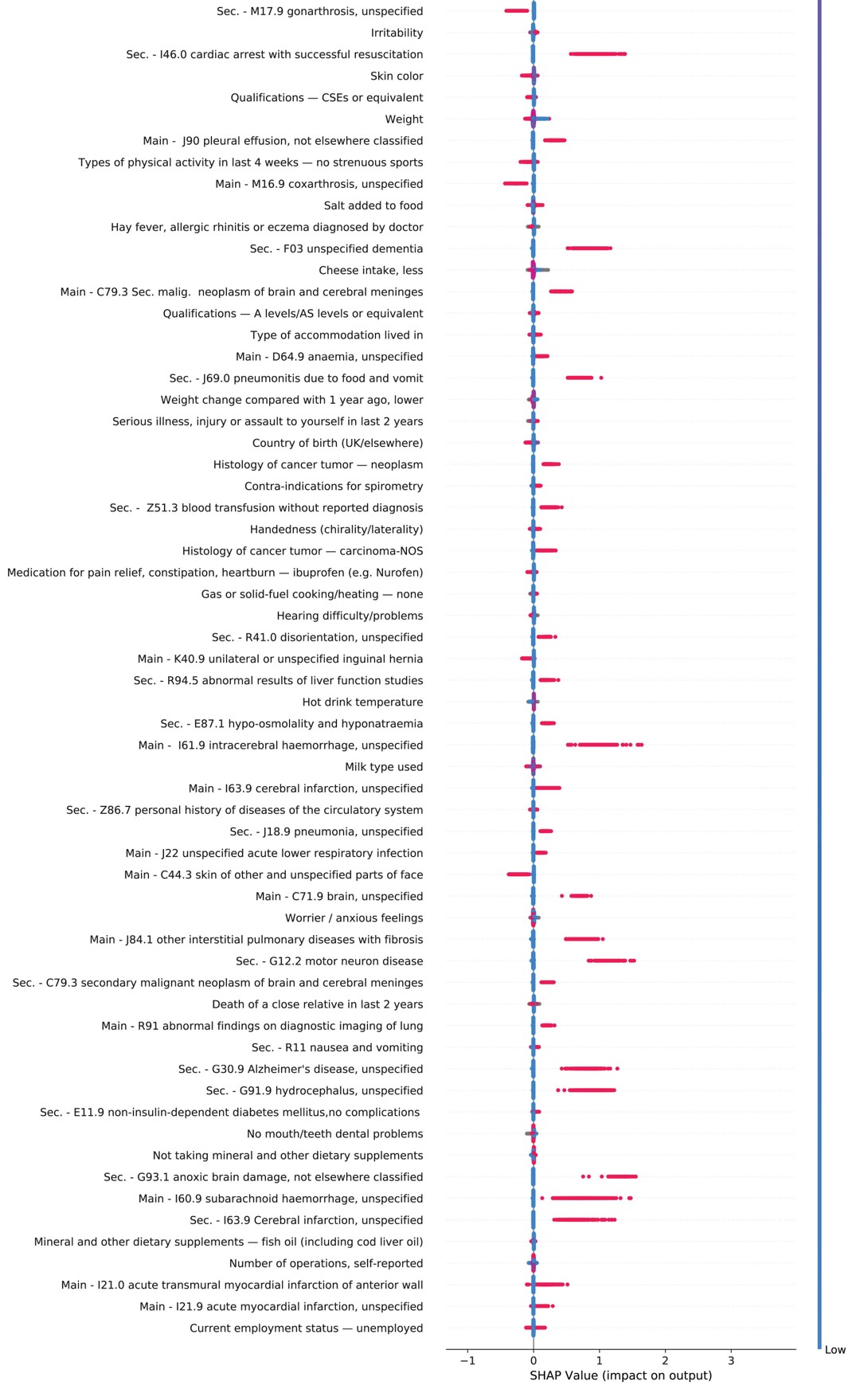

**Supplementary Figure 3**. SHAP (SHapley Additive exPlanation) variable importance for the 193 important predictors on all-cause mortality in the UK Biobank. Each point on the plot is a SHAP value for a predictor and a sample. The position on the y-axis is determined by the predictor and on the x-axis by the SHAP value. The colour gradient represents predictor values from low (blue) to high (red). Grey dots represent samples with missing information. International classification of diseases (ICD) codes are given for diagnoses. Predictor names are modified by adding additional text after a ‘—’ for some predictors to reflect how higher value(s) are coded. Abbreviations: A level, advanced level; AS level, advanced subsidiary level; CSE, certificate of secondary education; NOS, not otherwise specified; Sec., secondary diagnosis.

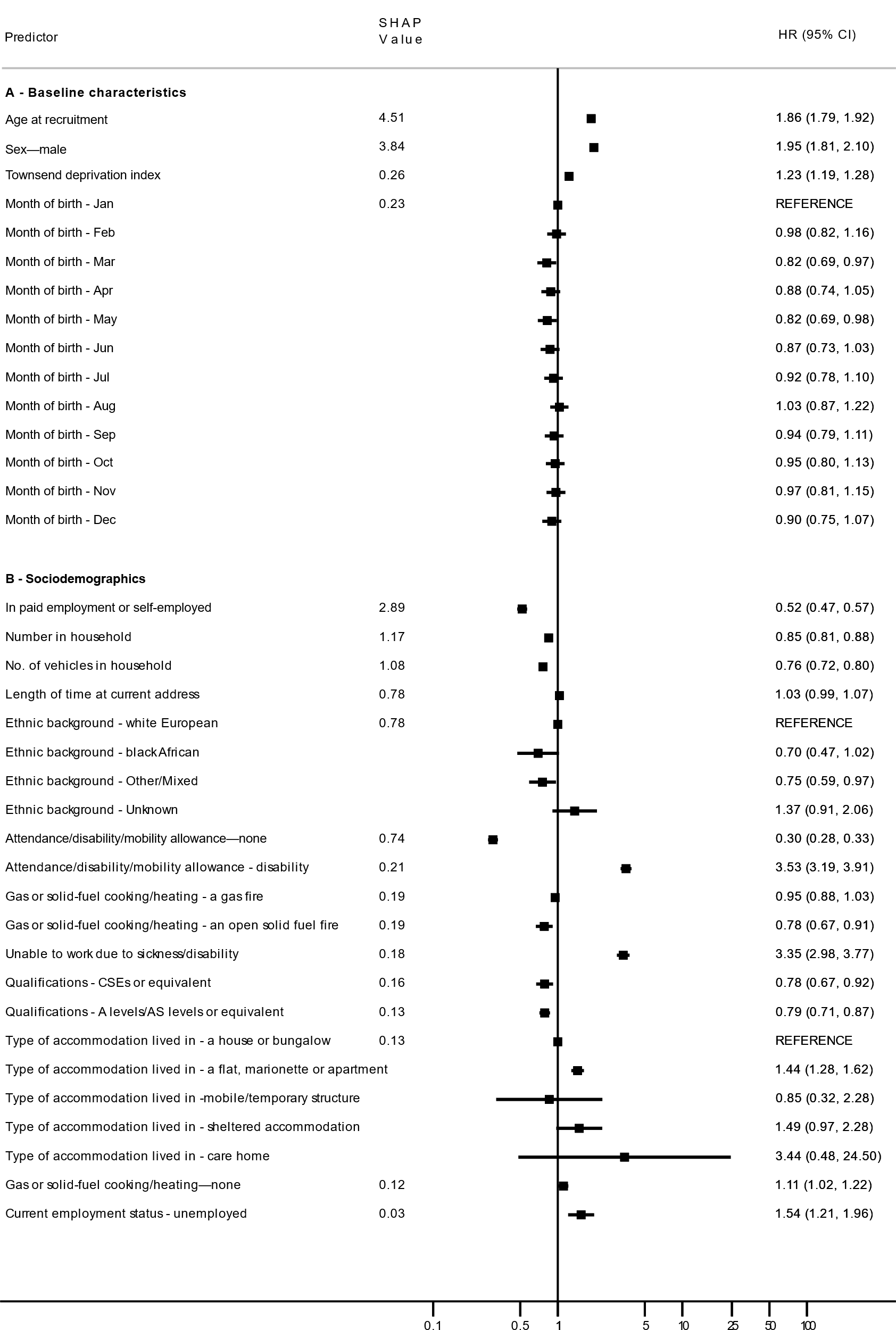

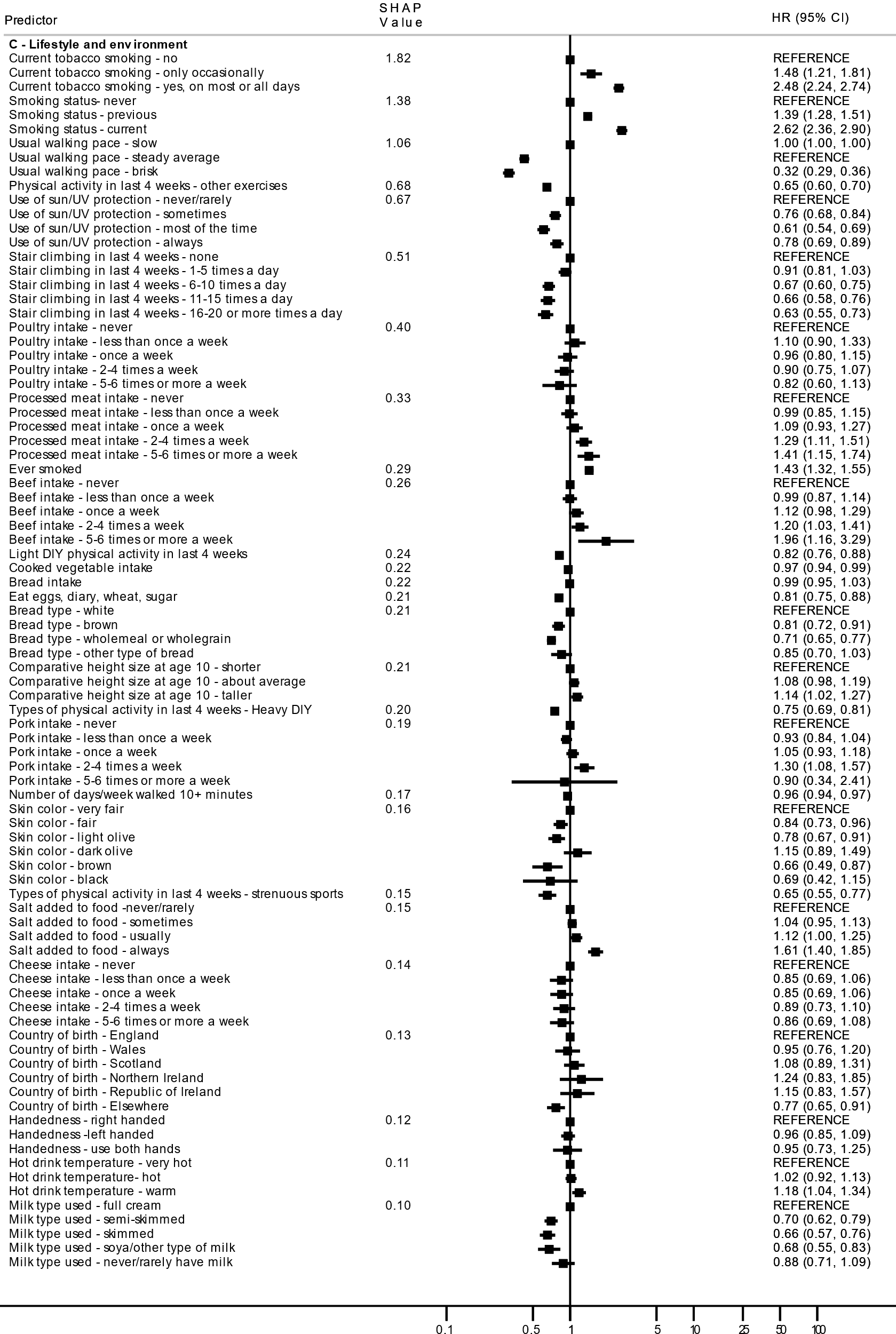

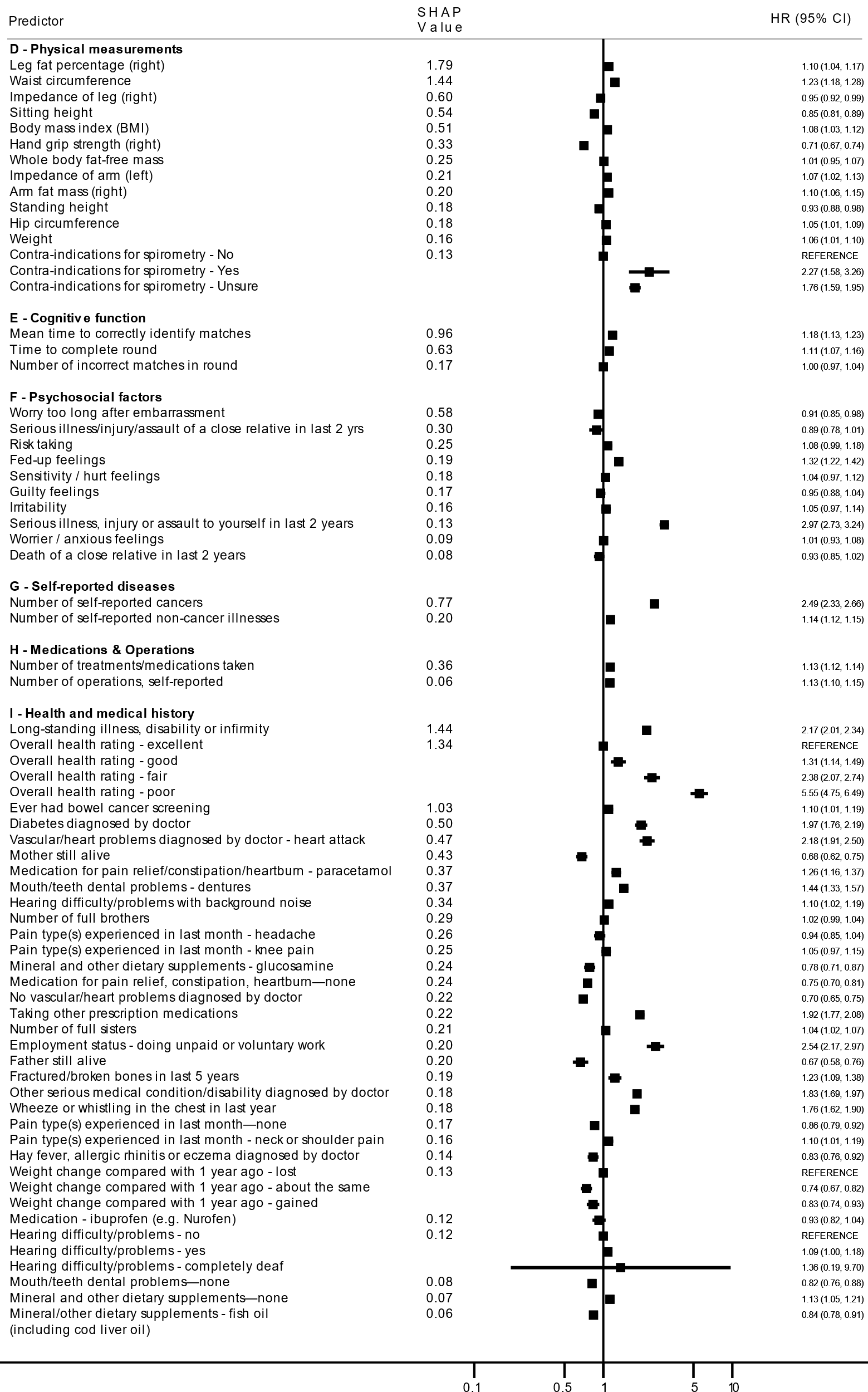

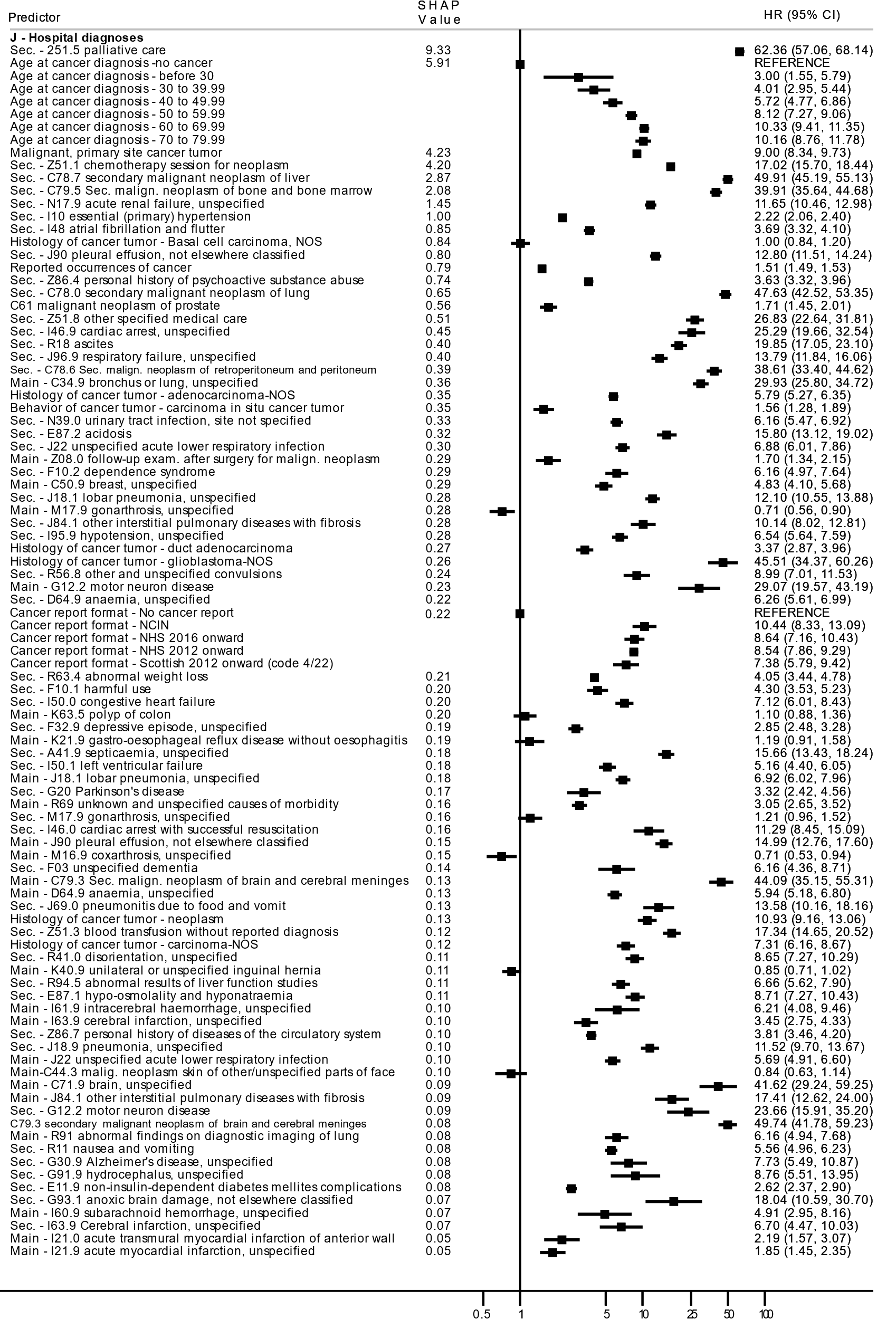

**Supplementary Figure 4.** Hazard ratio (HR) with 95% confidence interval and SHAP (SHapley Additive exPlanation) values arranged by predictor category for all the importance predictors (193 predictors). International classification of diseases (ICD) codes are given for diagnoses. Predictor names are modified by adding additional text after a ‘—’ for some predictors to reflect how higher value(s) are coded. Abbreviations: HR, hazard ratio; NOS, not otherwise specified; Sec., secondary diagnosis.

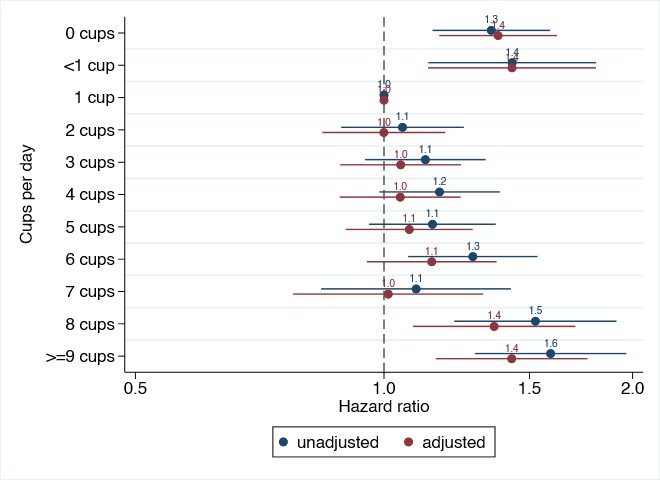

**Supplementary Figure 5.** Association of tea intake with all-cause mortality, unadjusted and adjusted for age, sex, Townsend deprivation index, assessment center and month of birth. Reference group is one cup per day. 95% confidence intervals are shown for hazard ratios using horizontal bars.

**References**

1	Kim SY, Kim S, Cho J*, et al.* A deep learning model for real-time mortality prediction in critically ill children. *Critical Care* 2019;**23**:279.

2	Meyer A, Zverinski D, Pfahringer B*, et al.* Machine learning for real-time prediction of complications in critical care: a retrospective study. *The Lancet Respiratory Medicine* 2018;**6**:905-14.

3	Mohamadlou H, Panchavati S, Calvert J*, et al.* Multicenter validation of a machine-learning algorithm for 48-h all-cause mortality prediction. *Health informatics journal* 2019:1460458219894494.

4	Weng SF, Vaz L, Qureshi N*, et al.* Prediction of premature all-cause mortality: A prospective general population cohort study comparing machine-learning and standard epidemiological approaches. *PloS one* 2019;**14**:e0214365.

5	Olson RS, La Cava W, Mustahsan Z*, et al.* Data-driven advice for applying machine learning to bioinformatics problems. *arXiv preprint arXiv:170805070* 2017.

6	Prokhorenkova L, Gusev G, Vorobev A*, et al.* CatBoost: unbiased boosting with categorical features. *Adv Neur In* 2018;**31**.

7	Dorogush AV, Ershov V, Gulin A. CatBoost: gradient boosting with categorical features support. *arXiv preprint arXiv:181011363* 2018.

8	Bentéjac C, Csörgő A, Martínez-Muñoz G. A comparative analysis of gradient boosting algorithms. *Artificial Intelligence Review* 2020:1-31.

9	Millard LA, Davies NM, Gaunt TR*, et al.* Software Application Profile: PHESANT: a tool for performing automated phenome scans in UK Biobank. *International Journal of Epidemiology* 2017.

10	Shapley LS. A value for n-person games. *Contributions to the Theory of Games* 1953;**2**:307-17.

11	Lundberg SM, Lee S-I. A unified approach to interpreting model predictions. *Advances in Neural Information Processing Systems* 2017:4765-74.

12	Lundberg SM, Lee S-I. Consistent feature attribution for tree ensembles. *arXiv preprint arXiv:170606060* 2017.
